# Worldwide diversity and geographic structure of killer-cell immunoglobulin-like receptor (KIR) gene content

**DOI:** 10.64898/2026.10.09.26365105

**Authors:** Alvaro Obeso

**Author notes:** Corresponding author Alvaro Obeso Fernandez, PhD, Helsinki Institute for Demography and Public Health, University of Helsinki, Snellmaninkatu 12, 00170 University of Helsinki Finland, GSM.

## Abstract

**Introduction:** Killer-cell immunoglobulin-like receptor (KIR) gene content shapes natural killer-cell responses and varies substantially among populations. We examined whether worldwide KIR diversity reflects regional structure, isolation by distance, serial founder effects, or latitudinal gradients.

**Data and methods:** Carrier frequencies for 11 variable KIR genes were obtained from the Allele Frequency Net Database. The primary analysis included 189 locally sampled, non-diaspora, non-mixed populations from 70 countries, comprising 25,469 individuals; 161 populations had complete 11-gene profiles. Diversity indices, ordination, clustering, permutation-based variance decomposition, Mantel tests, and regression models were used to characterize geographic structure. Serial founder effects were evaluated using four distance measures from East Africa, both globally and within the Old World. Models of latitude incorporated spatial autocorrelation. Prespecified sensitivity analyses evaluated alternative population sets, gene panels, geographic classifications, weighting schemes, and multivariate outlier exclusion.

**Results:** Geographic region explained 53.5% of variation in KIR profiles, substantially exceeding the permutation-null expectation of 6.9%, although multivariate dispersion differed among regions. Twenty-five of 28 pairwise regional comparisons remained significant after false-discovery-rate correction. Isolation by distance was modest (Mantel r=0.3003) and weakened after controlling for region (r=0.1557). KIR diversity did not decline with distance from East Africa globally but decreased strongly within the Old World (expected heterozygosity β=−0.0066 and standardized gene content β=−0.0789). B-content decreased nominally with absolute latitude, but latitude was collinear with region and the association survived neither spatial modelling nor sensitivity analyses. Regional differentiation remained robust across analytical specifications, whereas the Old World decline was carried by gene content once standard errors were clustered by country.

**Conclusion:** Worldwide KIR gene-content variation shows pronounced regional differentiation, modest isolation by distance, and no independent latitudinal gradient. Diversity declines with distance from East Africa across the Old World, supporting serial founder effects, but not once the Americas and Oceania are included.

## Introduction

Human natural killer (NK) cells recognise infected, transformed and otherwise altered cells through killer-cell immunoglobulin-like receptors (KIR), activating and inhibitory receptors binding predominantly human leukocyte antigen (HLA) class I ligands (1). Unlike the somatically generated antigen receptors of T and B lymphocytes, KIR diversity is germline encoded, comprising gene content and allelic variation that differs substantially among individuals and populations (2, 3). With cognate HLA variation, it has been associated with viral infection control (4), pregnancy outcomes through maternal–foetal KIR–HLA-C interactions (5), haematopoietic stem-cell transplantation outcomes (6) and susceptibility to autoimmune and inflammatory disease (7), making population-level KIR diversity relevant to human evolution and to clinical medicine.

The KIR locus, in the leukocyte receptor complex on chromosome 19q13.4, shows extensive allelic and structural variation, its haplotypes differing in gene number and composition (2, 3, 8–11). Group A haplotypes are relatively conserved and predominantly encode inhibitory receptors, whereas the more variable group B haplotypes carry additional activating genes, often alongside KIR2DL2 and KIR2DL5 (3, 8, 9). Centromeric and telomeric regions hold alternative A and B motifs with partly distinct evolutionary histories (3, 8). Framework genes being comparatively conserved, variation in gene presence is a major contributor to worldwide KIR diversity (9–11).

KIR gene content, haplotype and genotype frequencies vary markedly among geographic regions (9, 10, 12, 13). Covariation between activating KIR genes and their HLA class I ligands implicates demographic history and geographically varying selection (9), and balancing, purifying and positive selection have all been reported across KIR regions and populations (12). Whether this variation reflects isolation by distance, serial-founder effects or independent latitudinal gradients remains unclear.

Neutral human genetic diversity declines with geographic distance from Africa, reflecting successive founder events during human expansion (14, 15). Whether KIR gene-content diversity does so is uncertain, because demographic signals may be modified by selection on receptor–ligand combinations involved in immunity and reproduction (10, 12); an earlier global analysis found activating KIR genes and their HLA ligands decreasing with distance from East Africa but did not test overall KIR diversity (10). Geographic variation in pathogens and other selective pressures could likewise produce latitudinal differences in activating and inhibitory KIR profiles (12). Distinguishing such gradients from regional structure matters, because apparent distance or latitude associations may instead reflect differences among historically differentiated regions.

We therefore used Allele Frequency Net Database carrier frequencies to characterise worldwide diversity and geographic structure across 11 variable KIR genes, quantifying within-population diversity, partitioning multivariate variation among regions and countries, and testing whether KIR-profile dissimilarity increased with geographic distance, whether diversity declined with distance from East Africa as a serial-founder model predicts, and whether activating KIR content varied with latitude. Global, within-region and joint latitude–distance models were complemented by prespecified sensitivity analyses separating continuous gradients from regional differentiation.

## Data and methods

### Data source and study populations

Population-level KIR carrier frequencies were obtained from the Allele Frequency Net Database, accessed on September 18, 2026, using the KIR gene-frequency dataset (16). The extracted dataset comprised 270 populations and included population identifiers, geographic coordinates, sample sizes, genes reported, and carrier frequencies. Frequencies were converted from percentages to proportions. Genes not reported by a source study were treated as missing rather than absent because replacing unreported values with zero would artificially increase apparent differences among populations.

Eleven variable KIR genes were included in the analyses: KIR2DL1, KIR2DL2, KIR2DL3, KIR2DL5, KIR3DL1, KIR2DS1, KIR2DS2, KIR2DS3, KIR2DS4, KIR2DS5, and KIR3DS1. The framework genes KIR2DL4, KIR3DL2, and KIR3DL3 and the pseudogenes KIR2DP1 and KIR3DP1 were described separately but excluded from diversity and multivariate analyses because their carrier frequencies were near fixation.

Populations were classified by country, geographic region, broad geographic area and population history. A population was classified as diaspora when its label indicated ancestry geographically distinct from the sampling location, such as African Americans, Japanese sampled in Brazil, or Indians sampled in Singapore or South Africa. Populations labelled mixed, mixed-race, mixed-ancestry, Mestizo, Mestiza or Mestizos, or described in the database notes as locally admixed, were classified separately from geographically local populations. Because classification turned on recent relocation rather than ancestral origin, long-established populations were retained unless explicitly labelled diaspora or mixed ancestry; the Macedonian Romani, Mumbai Parsi and Chechens, resident in their sampling regions for centuries, were therefore treated as local, and excluding them changed regional variance negligibly (0.5349 vs 0.5342). Population characteristics, classifications, analysis-set assignments, and the database identifier and source publication of each population are given in Supplementary Table 1.

The primary analysis comprised geographically local populations reporting at least 9 of the 11 variable genes: 189 populations from 70 countries and 25,469 individuals, of which 161 had complete carrier-frequency data for all 11 genes and entered the distance-based multivariate analyses. Two sensitivity sets were defined: excluding populations of fewer than 50 individuals left 173 populations and 24,819 individuals, 146 with complete 11-gene profiles; including all populations reporting at least 9 variable genes, diaspora, mixed-ancestry and Mestizo among them, gave 236 populations from 78 countries and 31,469 individuals, 205 with complete profiles. Supplementary Figure 1 shows the study design and the populations retained and excluded at each stage.

### Geographic classification and distance measures

To examine geographic structure at different spatial scales, populations were assigned to 12 geographic regions: Sub-Saharan Africa, North Africa, Western Asia, Central Asia, South Asia, North-East Asia, South-East Asia, Europe, Australia, Oceania, North America, and South and Central America. A broader eight-area classification, combining related regions and separating Central from South America, was used in a sensitivity analysis. Because North-East Asia spanned 21.97° to 44.00° N and combined southern Chinese, Taiwanese, Yunnan minority, Tibetan and Uygur populations, a further partition split southern from northern East Asia at 30° N and placed Tibet and Xinjiang in their own groups.

Pairwise geographic distances were great-circle distances between population coordinates, computed with the haversine formula. Distance from East Africa was evaluated under four definitions: a waypoint-constrained route originating in Addis Ababa, and direct great-circle distances from Addis Ababa, Nairobi and Cairo. The waypoint route approximated terrestrial dispersal and avoided implausible transoceanic distances, running through Addis Ababa (9.03° N, 38.74° E), Cairo (30.04° N, 31.24° E), Istanbul (41.01° N, 28.98° E), Phnom Penh (11.55° N, 104.92° E), Anadyr (64.73° N, 177.51° E) and Prince Rupert (54.31° N, 130.32° W). Distances ran directly from Addis Ababa for sub-Saharan Africa, through Cairo for North Africa and Western Asia, through Istanbul for the remaining Old World, and through the full route for the Americas. Because waypoint distance was strongly collinear with region (η²=0.9788, against 0.8537–0.9292 for the three great-circle distances), their effects could not be fully separated; distance models were therefore fitted globally and within the Old World, where the four measures agreed more closely.

### KIR diversity and gene-content indices

Within-population diversity was quantified primarily using standardized gene content across the 11 variable genes, which requires no conversion of carrier frequencies or assumptions about allelic structure. Expected heterozygosity was evaluated secondarily. Because AFND reports carrier frequencies, the absence-allele frequency was estimated under Hardy–Weinberg equilibrium (HWE) using Bernstein’s formula q= √ (1-c), where c is the carrier frequency. The presence-allele frequency was calculated as p=1−q and expected heterozygosity as H_e_=2pq (10). This conversion is generally accurate but is less reliable for KIR2DS3 and KIR2DL5, which can occupy alternative positions within the locus (10). In addition, KIR2DL2/KIR2DL3 and KIR3DL1/KIR3DS1 are allelic pairs, meaning that the 11-gene mean counts two loci twice and introduces negative dependence (10). Accordingly, standardized gene content, which requires no carrier-to-allele conversion, was the primary diversity measure. Expected-heterozygosity and other gene-content analyses were supplemented by sensitivity analyses restricted to seven unambiguous marker genes. HWE could not be tested from population-level carrier frequencies.

The standardized gene-content index was calculated as the sum of available carrier frequencies multiplied by 11 and divided by the number of genes reported, thereby scaling each value to the full 11-gene panel. This population-level index should not be interpreted as an individual KIR gene count. Activating- and inhibitory-gene content indices were calculated as the mean carrier frequency of activating genes (KIR2DS1, KIR2DS2, KIR2DS3, KIR2DS4, KIR2DS5, and KIR3DS1) and inhibitory genes (KIR2DL1, KIR2DL2, KIR2DL3, KIR2DL5, and KIR3DL1), respectively. A- and B-content indices were similarly calculated from the mean carrier frequencies of A-associated genes (KIR2DL1, KIR2DL3, KIR3DL1, and KIR2DS4) and B-associated genes (KIR2DL2, KIR2DL5, KIR2DS1, KIR2DS2, KIR2DS3, KIR2DS5, and KIR3DS1), respectively. These population-level indices do not represent directly observed haplotype frequencies. Relative A- and B-content ratios, termed A-proportion and B-proportion, were calculated by dividing each content index by the mean carrier frequency across the full 11-gene panel. Worldwide distributions of the A- and B-content indices, framework genes, and pseudogenes are shown in Figure 1.

**Figure 1.**
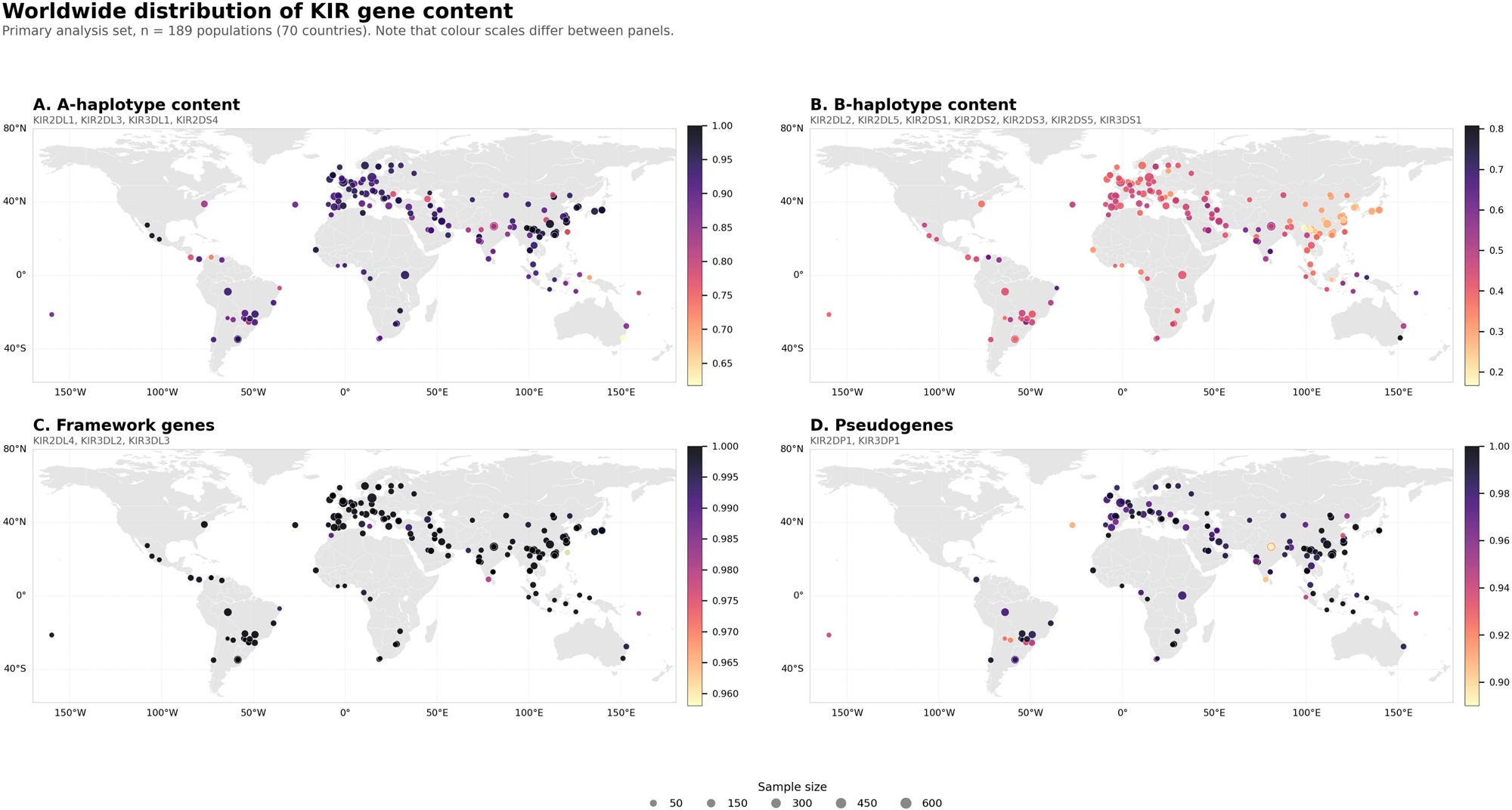
Worldwide distribution of KIR gene content in the primary analysis. **Caption:** Maps show population-level A-haplotype content index (A), B-haplotype content index (B), framework-gene carrier frequencies (C), and pseudogene carrier frequencies (D) among 189 geographically local populations from 70 countries. A-haplotype content index was calculated as the mean carrier frequency of KIR2DL1, KIR2DL3, KIR3DL1, and KIR2DS4. B-haplotype content index was calculated as the mean carrier frequency of KIR2DL2, KIR2DL5, KIR2DS1, KIR2DS2, KIR2DS3, KIR2DS5, and KIR3DS1. Framework-gene and pseudogene values were summarized descriptively. Point size represents sample size and color represents the corresponding population-level frequency index. Color scales differ among panels and should not be compared directly.

### Statistical analysis

Analyses followed a predefined hierarchy distinguishing the primary population set, the complete-profile subset, and the sensitivity analyses. Analyses of diversity, geographic distance and latitude used all populations with the variables required for the corresponding model, and multivariate analyses of complete gene-frequency profiles were restricted to the complete-profile subset. Log-transformed sample size was examined as a predictor of each gene-content measure before any geographic comparison. Log sample size was retained as a covariate in all subsequent regression models. All regression models were fitted by ordinary least squares, and the coefficients, standard errors and probability values reported in the text are from those fits. Because populations sampled within the same country are not statistically independent, country-clustered standard errors were additionally computed for the distance, latitude and joint models and are reported beside the ordinary-least-squares values. Model comparisons based on information criteria used unclustered fits throughout.

We first characterized overall KIR diversity and population structure. Expected heterozygosity and standardized gene content were compared among geographic regions in models that included log sample size. For complete profiles, Euclidean distance was the primary dissimilarity measure. Robustness to the choice of metric was evaluated using Manhattan, Bray-Curtis and standardized Euclidean distances, with agreement among matrices quantified by Mantel correlations. Population structure was visualized by principal-coordinate analysis. Hierarchical clustering used Ward’s minimum-variance method, and the number of clusters was selected by comparing mean silhouette widths for solutions containing 2 to 10 clusters. Agreement between data-derived clusters and geographic classifications was measured using the adjusted Rand index. Cluster stability was evaluated by resampling genes with replacement and calculating how frequently pairs of populations remained in the same cluster.

Geographic differentiation was assessed using distance-based permutation analyses that sequentially partitioned variation between region and country nested within region. Because the country term used 51 degrees of freedom across 63 countries, 38 represented by a single profile, each component was compared with a permutation distribution preserving the same unbalanced design; degrees of freedom and adjusted R^2^ are reported with the raw proportions. Pairwise tests included regions with at least three complete profiles, with p-values adjusted using the monotonic Benjamini–Hochberg false-discovery-rate procedure. Differences in within-region variability were evaluated using distances to regional centroids. Gene-specific differentiation among the 161 complete-profile populations was summarized using Nei’s G_ST_ = (H_T_-H_S_)/H_T_ where H_S_ is mean within-population expected heterozygosity and H_T_ is expected heterozygosity based on the mean allele frequency across populations. Presence-allele frequencies were inferred under HWE, populations were weighted equally, and no bias or small-sample correction was applied. Percentile confidence intervals were obtained from 1,000 bootstrap samples of populations drawn with replacement.

Having established the principal geographic structure, we tested whether it could be explained by continuous spatial or demographic gradients. Isolation by distance was assessed using the Mantel correlation between pairwise KIR-profile distances and great-circle geographic distances. A partial Mantel test additionally controlled for whether population pairs belonged to the same region. Because partial Mantel tests have recognized limitations, this analysis was treated as a secondary description rather than as evidence of a causal spatial process.

The serial-founder hypothesis was tested by modelling expected heterozygosity and standardized gene content as functions of distance from East Africa. The waypoint-constrained route was the prespecified primary definition, and the three great-circle definitions were examined as alternatives. To determine whether continuous distance or regional membership better described the observed variation, models containing sample size alone, distance, region, or both region and distance were compared using Akaike and Bayesian information criteria.

Latitudinal hypotheses were evaluated in the same regression framework. Absolute latitude was the primary predictor, because a pathogen-pressure rationale predicts variation with distance from the equator rather than with direction. Its associations with expected heterozygosity, standardized gene content, B-content, B-proportion and the activating and inhibitory indices were tested globally. Within-region models were fitted for regions represented by at least 10 populations, with probability values adjusted separately by outcome. Signed latitude was examined as a prespecified secondary predictor of directional north-south variation. Hemisphere-specific effects were assessed by including absolute latitude, hemisphere and their interaction. Potential nonlinearity was evaluated by adding a quadratic absolute-latitude term and comparing linear and quadratic specifications using information criteria and nested-model tests. Joint models included both absolute latitude and waypoint-constrained distance to examine whether either predictor retained an independent association.

Because nearby populations may remain similar after accounting for the specified predictors, residual spatial autocorrelation was evaluated using Moran’s I on a row-standardized eight-nearest-neighbour network constructed from population coordinates. Moran’s I was calculated for residuals from the primary B-content latitude model, the expected-heterozygosity distance model, and the corresponding joint models. Where autocorrelation was present, the latitude and distance models were refitted as spatial models: generalised least squares with an exponential correlation structure whose range and nugget were estimated by maximum likelihood, and ordinary least squares after Moran eigenvector filtering, with eigenvectors selected to minimise residual autocorrelation.

Robustness was assessed through prespecified sensitivity analyses. Primary analyses were repeated after restricting the dataset to populations with ≥50 individuals; including eligible diaspora, mixed-ancestry, and Mestizo populations; replacing the 12-region scheme with the broader eight-area classification; and sequentially omitting each region. Additional analyses used a finer East Asian partition, collapsed populations sharing a sampling location, and adjusted the regional model for publication year and typing method, both taken from the database record of each source study. Gene-content analyses were also restricted to complete profiles and to seven unambiguous A- or B-associated genes, excluding the allelic pairs KIR2DL2/KIR2DL3 and KIR3DL1/KIR3DS1. To address unequal precision arising from sample sizes of 19–690, regression models were weighted by sample size and, separately, by inverse binomial sampling variance. Because distance-based permutation analysis does not accommodate case weights, the regional model was repeated at progressively stricter sample-size thresholds. Sparse regional representation was assessed by excluding Central Asia, North America, Australia, and North Africa, each represented by fewer than three populations. A separate 10-gene analysis excluded KIR2DL5, recalculated expected heterozygosity, standardized gene content, B-content, B-proportion, and the inhibitory index, and repeated the regional differentiation, isolation-by-distance, latitude, distance, and joint models. Regression models in this analysis used all 189 primary populations, and the multivariate analyses used the 175 populations with complete 10-gene profiles. Five were excluded from that subset because the gene they did not report was not KIR2DL5.

Multivariate outliers were identified among complete profiles using Mahalanobis distance, with probabilities obtained from a chi-square distribution on 11 degrees of freedom and adjusted using the Benjamini-Hochberg procedure. Populations with an adjusted p < 0.05 were classified as outliers. They were retained in all primary analyses and excluded only in sensitivity analyses, in which the regional, isolation-by-distance, latitude, distance and joint models were repeated. Primary and sensitivity estimates were compared using sample size, regional R², Mantel correlation, regression coefficients, standard errors and probability values.

Simulation-based power analyses estimated the probability of detecting prespecified effects of absolute latitude on B-content and of waypoint-constrained distance on expected heterozygosity, conditional on the observed sample size, predictor distributions, association with log sample size and residual variance.

All analyses were performed in Python using pandas, NumPy, SciPy, scikit-learn, statsmodels, Matplotlib, Seaborn, and python-docx. Permutation, bootstrap, and simulation procedures used a fixed random seed of 20260923. The primary permutation tests and the pairwise regional comparisons used 9,999 permutations, secondary tests used 999 permutations, cluster stability and gene-specific confidence intervals used 1,000 bootstrap replicates and power analyses used 1,000 simulations per effect size. All probability values were two-sided. FDR adjusted p<0.05 was considered statistically significant where multiple comparisons were performed.

## Results

### Sampling effort and regional diversity

The primary analysis comprised 189 geographically local populations from 70 countries, representing 25,469 individuals, of which 161 had complete data for all 11 variable genes. Coverage was uneven across regions: Europe contributed 52 populations and North-East Asia 35, whereas Central Asia and North America contributed one each and Australia and North Africa two each, which constrains the precision of the regional estimates reported below.

Expected heterozygosity, the standardized gene-content index, and the inhibitory index showed no material association with sample size. B-content decreased weakly with increasing sample size (β=−0.0282, p=0.0185), whereas the activating-index association did not reach statistical significance (p=0.0647). Log-transformed sample size was therefore retained as a covariate in subsequent regression models (Supplementary Table 2). Geographic region was strongly associated with both expected heterozygosity and the standardized gene-content index in the primary and sensitivity analyses (all p<0.0001) (Supplementary Table 3).

### Geographic structure of KIR profiles

Geographic region accounted for 53.5% of total variation in complete KIR profiles, country within region for a further 18.8%, leaving 27.7% as residual population-level variation (Table 1). Against a permutation null with the same unbalanced design, the regional component lay far above chance (null mean 6.9%, 95th percentile 10.1%; adjusted R² = 0.5005, p = 0.0005), whereas the country-within-region component fell below its own null (null mean 31.8%, p = 1.0000), reflecting the 51 degrees of freedom it consumes rather than explained variation. Regional dispersion also differed significantly (F = 8.35, permutation p = 0.0001), so the regional effect combines differences in profile location with unequal within-region heterogeneity. Twenty-five of 28 eligible pairwise regional comparisons remained significant after false-discovery-rate correction, the strongest being North-East Asia against Sub-Saharan Africa (R² = 0.6301, p = 0.0001); North-East Asia featured in four of the five strongest comparisons (Supplementary Figure 2 with estimates in Supplementary Table 4).

**Table 1.** Variance decomposition, spatial association and primary regression models for the 189-population primary analysis set. Variance components represent the proportion of total variation in KIR genefrequency profiles attributable to geographic region, country nested within region, and differences among populations. Differences in multivariate dispersion were assessed across geographic regions. Mantel tests evaluated the association between KIR-profile dissimilarity and geographic distance, with the partial Mantel test controlling for geographic region. Regression coefficients (β) represent the change in B-content per degree of absolute latitude and the change in expected heterozygosity per 1,000 km of waypoint distance. **Abbreviations**: KIR: killer-cell immunoglobulin-like receptor; β: regression coefficient; SE: standard error; F: F-statistic; r: correlation coefficient; p: p-value. Dashes indicate not applicable.

|  | Estimate | Test statistic | p |
| --- | --- | --- | --- |
| *Variance decomposition of KIR profiles* |  |  |  |
| Geographic region | 53.5% of total variation | F = 15.58 | 0.0001 |
| Country within region | 18.8% of total variation | — | — |
| Residual (among populations) | 27.7% of total variation | — | — |
| Multivariate dispersion between regions | — | F = 8.35 | 0.0001 |
| *Spatial association* |  |  |  |
| Mantel, KIR profile vs geographic distance | r = 0.3003 | — | 0.0010 |
| Partial Mantel, region controlled | r = 0.1557 | — | 0.0021 |
| *Primary regression models* |  |  |  |
| B-content on absolute latitude | $\beta = -0.0010$ per degree | SE = 0.0005 | 0.0419 |
| Expected heterozygosity on waypoint distance | $\beta = 0.0008$ per 1,000 km | SE = 0.0005 | 0.1478 |

The first two principal-coordinate axes accounted for 74.38% of variation among complete profiles and showed broad geographic structuring with regional overlap (Figure 2). Results were consistent across distance measures, with correlations among Euclidean, Manhattan, Bray–Curtis, and standardized Euclidean matrices of 0.9646–0.9898 (Supplementary Table 5).

**Figure 2.**
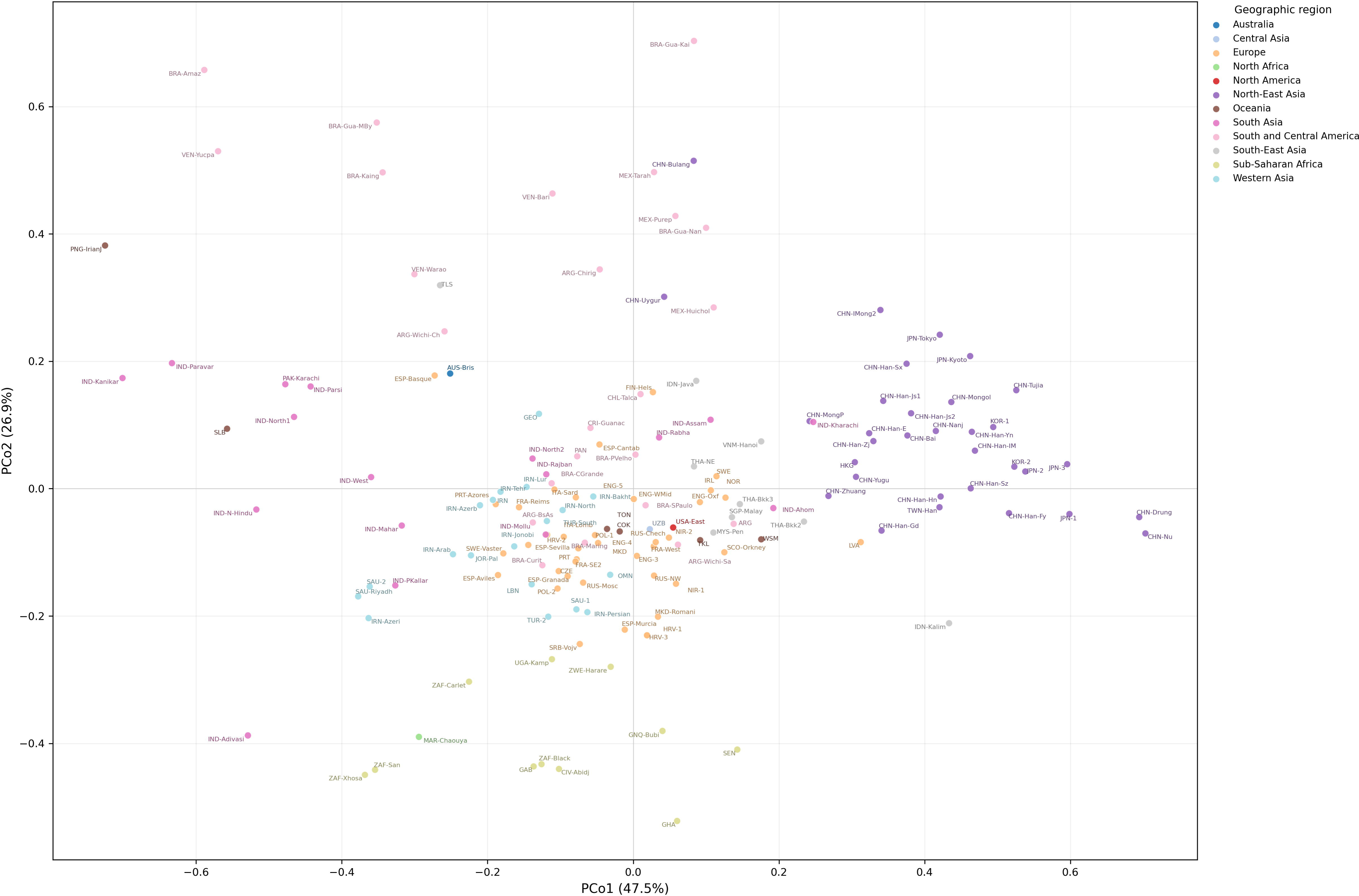
Geographic structure of worldwide KIR carrier-frequency profiles. Principal-coordinate analysis of Euclidean distances among the 161 populations with complete carrier-frequency profiles for all 11 variable KIR genes. Each point represents one population, colors indicate geographic region, and labels identify populations using the abbreviations defined in Supplementary Table 1. The first and second principal-coordinate axes explained 47.5% and 26.9% of the total variation, respectively. **Abbreviations:** PCo1: first principal-coordinate axis; PCo2: second principal-coordinate axis.

Mean silhouette width was highest for the seven-cluster solution. However, concordance with geographic region was modest (adjusted Rand index=0.3315), median gene-resampling stability was 0.6519, and adjusted Rand indices varied little among solutions with four or more clusters (Supplementary Figure 3 with estimates in Supplementary Table 6).

Gene-specific differentiation among the 161 complete-profile populations varied across the 11 loci. The highest G_ST_ estimates were observed for KIR2DS4 (0.1065), KIR2DL3 (0.1045) and KIR2DL1 (0.0972), and the lowest for KIR2DL5, KIR2DS1 and KIR2DS3. Bootstrap confidence intervals and complete results are given in Supplementary Figure 4 with estimates in Supplementary Table 7.

### Isolation by distance and serial-founder analyses

KIR-profile dissimilarity was modestly correlated with geographic distance (Mantel r = 0.3003, p = 0.0010). The association weakened after controlling for whether population pairs belonged to the same region (partial Mantel r = 0.1557, p = 0.0021) (Table 1).

Under the prespecified waypoint-constrained route, neither expected heterozygosity nor standardized gene content declined with distance from East Africa; both coefficients were positive and non-significant (β = 0.0008 per 1,000 km, p = 0.1478 and β = 0.0018, p = 0.7006, respectively). All three great-circle definitions instead gave negative coefficients, significant for standardized gene content from every origin (β = −0.0298 to −0.0470, p = 0.0096 to p = 0.0008) and for expected heterozygosity from Nairobi only (β = −0.0035, p = 0.0267). Among the 155 populations outside the Americas, Oceania and Australia, both measures declined steeply with waypoint-constrained distance (expected heterozygosity β = −0.0066 per 1,000 km, p = 0.0001; standardized gene content β = −0.0789, p < 0.0001), the great-circle definitions agreed in direction and significance, and a formal interaction confirmed that the slope differed between the Old World and the remaining populations (p = 0.0085 and p = 0.0005, respectively). Diversity therefore declined with distance from East Africa across the Old World but not worldwide, the global null arising from the 34 American, Oceanian and Australian populations, which combine the largest dispersal distances with high KIR diversity (Supplementary Figure 5 with estimates in Supplementary Tables 8 and 9). Clustering standard errors by country left the Old World gene-content decline and its interaction significant (p = 0.0092 and p = 0.0426) but not their expected-heterozygosity equivalents (p = 0.0823 and p = 0.1428), and left no great-circle association significant across all populations.

### Latitude and spatial-autocorrelation analyses

B-content showed a weak inverse association with absolute latitude in the primary model (β = −0.0010 per degree, p = 0.0419), with a similar pattern for B-proportion (β = −0.0012, p = 0.0298) (Supplementary Table 10). No within-region association remained significant after FDR correction, the strongest being a negative association in Europe (β = −0.0023 per degree, raw p = 0.0112, FDR p = 0.0784) (Supplementary Figure 6 with estimates in Supplementary Table 11), and the association was attenuated after joint adjustment for waypoint-constrained distance from East Africa (β = −0.0009, p = 0.1078) (Figure 3 with estimates in Supplementary Table 12).

**Figure 3.**
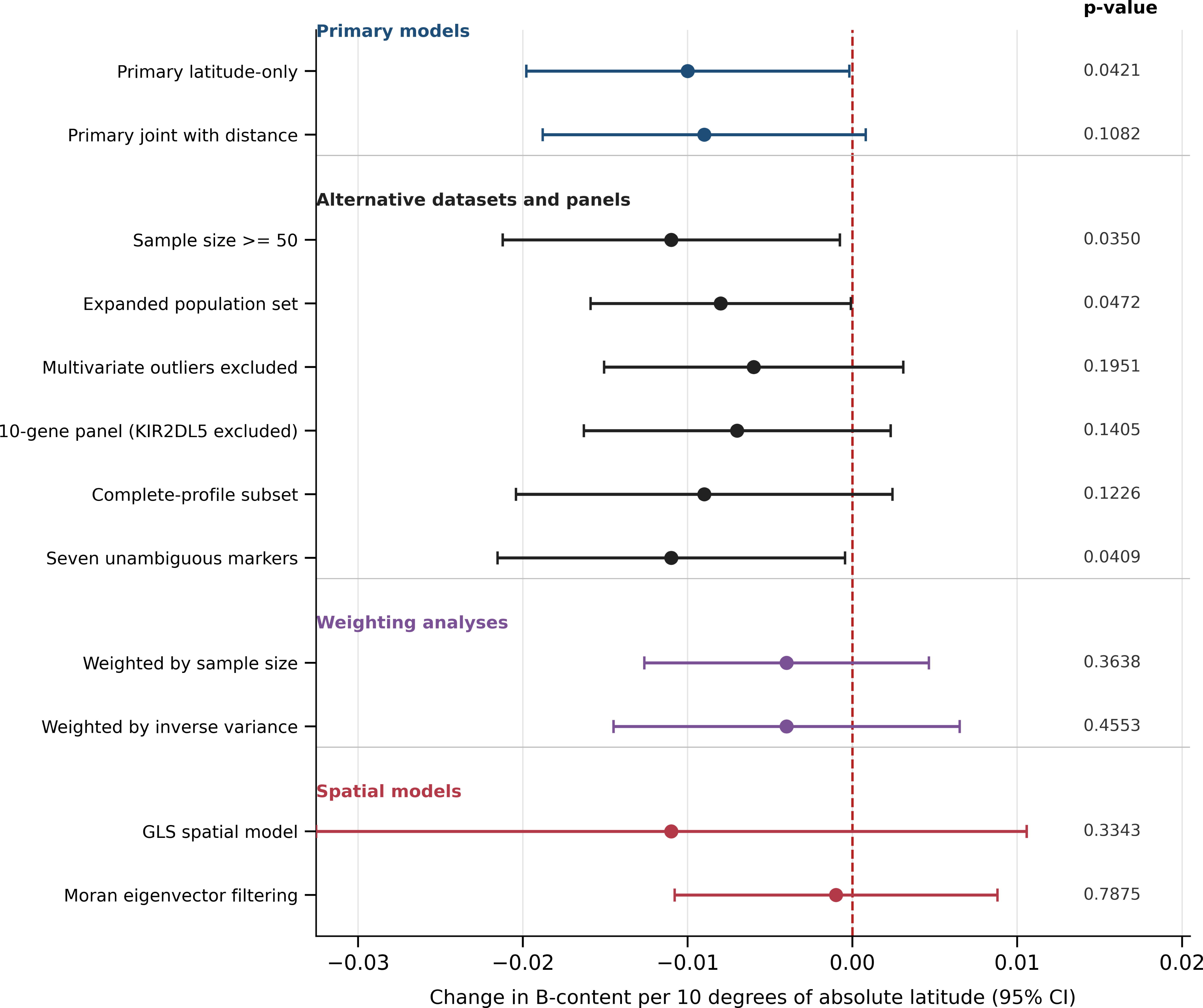
Sensitivity of the association between KIR B-content and absolute latitude. **Caption:** Points represent estimated changes in the B-content index per 10° increase in absolute latitude, and horizontal lines indicate 95% confidence intervals. Latitude-only models included absolute latitude and log-transformed sample size; joint models additionally included waypoint-constrained distance from East Africa. Estimates are shown for the primary 11-gene latitude-only and joint models; for the sample-size-restricted analysis, the expanded population set, the analysis excluding FDR-defined multivariate outliers, the 10-gene panel excluding KIR2DL5, the complete-profile subset and the seven unambiguous markers; for the two precision-weighted models; and for the two spatial models. The vertical dashed line indicates no association. Nominal two-sided p-values are shown; confidence intervals crossing zero indicate that the corresponding association was not statistically significant at p<0.05. **Abbreviations:** CI: confidence intervals.

The latitude estimate was small and imprecise and below the smallest effect detectable with 80% power, which lies between 0.0010 and 0.0015 per degree (Supplementary Table 13). Weighting by sample size or inverse sampling variance removed the association (β=−0.0004, p=0.3638 and β=−0.0004, p=0.4553, respectively) (Supplementary Table 14). Absolute latitude was strongly associated with region (η²=0.7627): all 32 populations above 45° were European, and none in the Southern Hemisphere exceeded 35°, making the hemisphere interaction unidentifiable across the latitude range of the northern slope. Spatial models also removed the association (generalised least squares: β=−0.0011, p=0.3343; Moran eigenvector filtering: β=−0.0001, p=0.7875), with filtering reducing residual Moran’s I from 0.4387 to 0.0001 (Supplementary Table 15). Clustering standard errors by country also removed the absolute-latitude association (p = 0.1143), whereas the signed-latitude associations were retained (Supplementary Table 10).

Signed latitude was negatively associated with B-content (β=−0.0009, p=0.0021) and B-proportion (β=−0.0011, p=0.0006; Supplementary Table 10). Hemisphere interactions and quadratic latitude terms were not statistically significant, although the uneven hemispheric distribution limited estimation of hemisphere-specific slopes (Supplementary Figure 7 with estimates in Supplementary Table 16). Residual spatial autocorrelation persisted in the absolute-latitude, distance, and joint models (Moran’s I=0.4387–0.4564; permutation p=0.0010) (Supplementary Figure 8 with estimates in Supplementary Table 17).

### Sensitivity analyses

Regional differentiation and isolation by distance persisted across the alternative analyses (Table 2). Regional R² ranged from 0.5343 to 0.5802 on restricting to populations with ≥50 individuals, excluding KIR2DL5, or removing multivariate outliers (Table 2; outlier screen in Supplementary Table 18), and remained significant under the broader area classification and the leave-one-region-out analyses. Omitting North-East Asia reduced it from 0.5349 to 0.4124, whereas omitting any other region changed it by less than 0.05 (Supplementary Figure 9 with estimates in Supplementary Table 19). It fell to 0.4260 on including diaspora, mixed-ancestry and Mestizo populations, rose to 0.5503 under the finer 14-group East Asian partition, and was 0.5420 after collapsing the 161 profiles into 145 distinct sampling locations, where the Mantel correlation changed from 0.3003 to 0.2992. Excluding the four regions represented by fewer than three populations gave 0.5181, rising to 0.5363 at a sample-size threshold of 50 and 0.5442 at 100 (Supplementary Table 22). Typing method and publication year were recovered for 178 of the 189 populations and 151 of the 161 complete profiles; adjusting for both left the regional component unchanged or slightly stronger, and neither publication year (p = 0.7991 and 0.8675) nor typing platform (p = 0.1544 and 0.1916) was associated with either measure (Supplementary Table 20). Carrier frequencies differed by typing method for KIR2DL2, KIR2DL3 and KIR2DS2 but not for KIR2DS4, KIR2DL5, KIR2DS3 or KIR2DS5, the genes for which method-dependent false negatives have been reported (Supplementary Table 21).

**Table 2.** Headline outcomes across the prespecified sensitivity analyses. Results from the primary analysis are compared with those obtained after restricting the dataset to populations with at least 50 individuals, including diaspora, mixed, and Mestizo populations, excluding multivariate outliers, and removing KIR2DL5 from the gene panel. Profiles indicate populations with complete data for all genes included in the corresponding panel. Regional R^2^ represents the proportion of variation in KIR genefrequency profiles explained by geographic region. Mantel *r* quantifies the association between KIR-profile dissimilarity and geographic distance. Latitude β represents the change in B-content per degree of absolute latitude, whereas distance β represents the change in expected heterozygosity per 1,000 km of waypoint distance. **Abbreviations**: KIR: killer-cell immunoglobulin-like receptor; R^2^: coefficient of determination; *r*: correlation coefficient; β: regression coefficient.

| Analysis | Populations | Profiles | Regional<br>$R^2$ | Mantel<br>$r$ | Latitude<br>$\beta$ | $p$ | Distance<br>$\beta$ | $p$ |
| --- | --- | --- | --- | --- | --- | --- | --- | --- |
| Primary analysis<br>(11 genes) | 189 | 161 | 0.5349 | 0.3003 | -0.0010 | 0.0419 | 0.0008 | 0.1478 |
| Sample size $\geq 50$ | 173 | 146 | 0.5551 | 0.3362 | -0.0011 | 0.0350 | 0.0006 | 0.2722 |
| Expanded set<br>(diaspora, mixed,<br>Mestizo) | 236 | 205 | 0.4260 | 0.2051 | -0.0008 | 0.0472 | 0.0005 | 0.1565 |
| Multivariate outliers<br>excluded | 175 | 147 | 0.5802 | 0.3203 | -0.0006 | 0.1951 | 0.0006 | 0.2960 |
| 10-gene panel<br>(KIR2DL5 excluded) | 189 | 175 | 0.5343 | 0.3212 | -0.0007 | 0.1405 | 0.0007 | 0.1721 |

The inverse absolute latitude association was significant in populations with ≥50 individuals (β=−0.0011, p=0.0350), the expanded dataset (β=−0.0008, p=0.0472), and the seven-marker analysis (β=−0.0011, p=0.0409). It was not significant after outlier removal (β=−0.0006, p=0.1951), exclusion of KIR2DL5 (β=−0.0007, p=0.1405), or restriction to complete profiles (β=−0.0009, p=0.1226) (Figure 3 with estimates in Table 2).

## Discussion

In this worldwide analysis of KIR carrier-frequency profiles, geographic region was the predominant correlate of population variation, whereas geographic distance accounted for a smaller component. KIR diversity declined with distance from East Africa within the Old World but not in the Americas, Oceania, and Australia. No latitudinal association could be separated from regional and spatial structure.

The pronounced regional differentiation is consistent with previous evidence that KIR gene-content, haplotype, and genotype distributions vary substantially among populations because of demographic history, migration, founder effects, and geographically varying selection (9,10,12,13,17). High-resolution studies also show considerable allelic and haplotypic diversity within populations that may not be captured by broad regional classifications (18,19). For example, extensive KIR and HLA class I diversity in an Egyptian population illustrates how local demographic history can produce patterns that differ from continental groupings (19).

Geographic region accounted for more than half of the variation in carrier-frequency profiles. Nevertheless, regional overlap in the ordination and the modest correspondence between data-derived clusters and geographic categories showed substantial population-level heterogeneity. This combination of regional structure and local variation is compatible with the effects of migration, drift, gene flow, and geographically varying selection involving KIR and HLA class I (9,12,18–21). Competing selective pressures related to infection and reproduction may also maintain A- and B-like haplotypes within populations rather than favouring a single KIR configuration (1,20,21).

KIR-profile dissimilarity increased modestly with geographic distance but weakened after controlling for region, indicating that much of the pattern reflected regional differentiation rather than a uniform global cline. Within the Old World, KIR diversity declined with distance from East Africa, consistent with the serial-founder model of human expansion (14,15). The decline did not extend to the Americas, Oceania and Australia, where populations combined long dispersal distances with relatively high KIR diversity. There, founder effects may have been offset by population-specific selection or the retention of distinctive haplotype combinations, as the high KIR diversity reported in Amerindian populations suggests; KIR haplotypes vary extensively in gene content, structure, copy number and alleles, and KIR and HLA class I show population-specific selection and coevolution (3,9,11,12,18,21). However, the waypoint route was assigned by region, so distance from East Africa was strongly collinear with it and these models describe a broad geographic gradient without isolating a dispersal process from regional structure.

B-content showed a modest inverse association with latitude, but this association weakened after adjustment for geographic distance, was absent within regions, and was sensitive to outlier exclusion, removal of KIR2DL5, population weighting, and spatial modelling. Absolute latitude was also strongly associated with region: all populations above 45° absolute latitude were European, and none in the Southern Hemisphere exceeded 35°. Consequently, hemisphere-specific slopes could not be reliably estimated across the relevant latitude range. Signed latitude also fitted the data better than absolute latitude, which is inconsistent with a simple equatorial pathogen-pressure hypothesis and more suggestive of a north–south ancestry gradient.

Geographic variation in pathogen exposure and reproductive pressures may still contribute to the distribution of A- and B-associated genes, but these pressures are unlikely to follow a uniform global cline. A- and B-like haplotypes have complementary roles in infection control and reproductive success, and their coexistence may be maintained by competing selective pressures (1,20,23). Selection may also act on specific KIR genes, alleles, expression levels, or KIR–HLA combinations rather than on average B-content (12,18,21,24). The latitude association should therefore be considered exploratory.

Residual spatial autocorrelation further limited the latitude analysis. Significant Moran’s I values after adjustment for latitude and distance showed that neighbouring populations remained more similar than predicted by the non-spatial models. Spatial modelling removed the latitude association, indicating that the nominal non-spatial result should not be interpreted independently of its geographic context.

The contribution of this study lies not in the regional stratification itself, which overlaps with previous work using the same database in 153 of the 189 populations analysed here (13), but in the hypothesis tests and robustness analyses applied to it. These include the joint evaluation of regional, distance, and latitude effects, the identification of a serial-founder pattern within the Old World but not globally, and the finding that the latitudinal association did not persist after accounting for spatial structure. The main regional and isolation-by-distance findings held across the prespecified sensitivity analyses, although both were weaker once diaspora, mixed-ancestry and Mestizo populations were included, and the serial-founder signal was carried by standardized gene content rather than by expected heterozygosity once standard errors were clustered by country. The use of continuous carrier-frequency profiles also retained information that would have been lost by reducing populations to categorical genotypes.

Nevertheless, several limitations apply. First, the analysis relied on aggregated carrier-frequency data from heterogeneous sources, with uneven geographic representation and variation in sample size, sampling strategy and genotyping methodology; underrepresentation of populations from Africa, Latin America and other regions is a broader limitation of KIR and HLA research that may restrict evolutionary inference and the transferability of clinical associations (19,25). Second, carrier frequencies carry limited information on phased haplotypes, allelic variation, copy number, structural variants and individual-level KIR–HLA combinations; earlier work emphasized the difficulty of inferring haplotypes and selection from marginal gene-frequency data alone (10), and sequencing-based approaches illustrate the technical challenges of homologous genes, gene absence, hybrid loci, duplications and copy-number variation (11,26). Third, HLA class I ligand frequencies could not be accounted for; because KIR function depends on the presence, allelic identity and expression of those ligands, identical gene-content profiles may differ functionally across populations with different HLA backgrounds (9,18,19,21). Fourth, latitude and geographic distance are broad spatial proxies that cannot distinguish among correlated demographic, climatic, pathogenic, nutritional and reproductive pressures, and the cross-sectional ecological design identifies geographic patterns without attributing them to specific evolutionary mechanisms or extending them to individuals. Finally, KIR and HLA polymorphisms have been associated with infectious disease, cancer, inflammatory and autoimmune disorders, reproductive outcomes and transplantation, but such effects generally depend on specific receptor–ligand combinations and clinical contexts (4–8,21,27), so high-resolution, well-powered studies in diverse populations are needed before these population-level frequency patterns can inform clinical prediction or donor selection.

In conclusion, worldwide KIR gene-content variation is characterized by pronounced regional differentiation and a decline in diversity with distance from East Africa within the Old World, but not globally. No latitudinal association was separable from regional and spatial structure. Future studies integrating high-resolution KIR haplotypes, alleles, copy number, and HLA ligand variation with demographic, environmental, and clinical data are needed to clarify the evolutionary and medical significance of worldwide KIR diversity.

## Ethics statement

This study used publicly available, aggregated population-level data from the Allele Frequency Net Database and did not involve the recruitment of participants, access to individual-level identifiable information, or collection of new biological samples. Therefore, institutional ethics approval and informed consent were not required.

## Author contributions

AO conceived and designed the study, collected, curated, and processed the data, developed the analytical methodology, performed all statistical analyses, interpreted the findings, and prepared the tables, figures, and supplementary material. AO wrote the original manuscript. All authors contributed to the improvement of the manuscript through critical review and revision. All authors read and approved the final version of the manuscript.

## Data Availability

The population-level KIR carrier-frequency data analysed in this study are publicly available from the Allele Frequency Net Database at https://www.allelefrequencies.net. The analysis code are available from the corresponding author upon reasonable request.

