## Supplementary material for "Worldwide diversity and geographic structure of killer-cell immunoglobulin-like receptor (KIR) gene content"

**Supplementary Figure 1.** Study design and analytical framework.
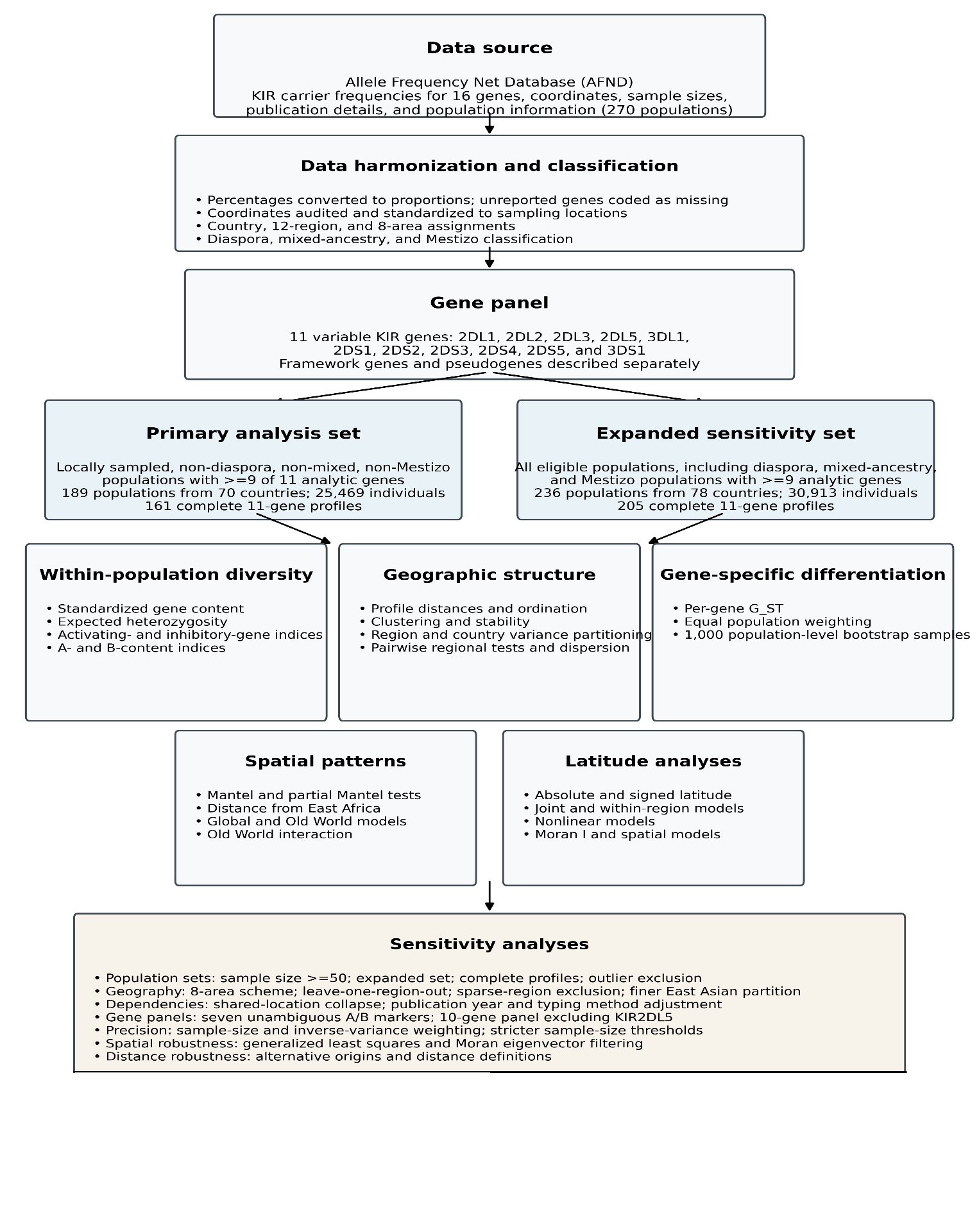

**Caption:** Population-level KIR carrier-frequency data and associated metadata were obtained from the Allele Frequency Net Database. The primary analysis included locally sampled populations not classified as diaspora, mixed-ancestry, or Mestizo, whereas the expanded sensitivity set included all eligible populations. Eleven variable KIR genes were used to quantify within-population diversity, geographic structure, isolation by distance, serial-founder patterns, and latitude associations. Robustness analyses evaluated alternative population sets, gene panels, geographic classifications, exclusion criteria, weighting schemes, and spatial models. Of 270 population records, 34 reporting fewer than 9 of the 11 variable genes were excluded. Of the remaining 236 populations, 47 classified as diaspora, mixed-ancestry, or Mestizo were reserved for sensitivity analysis, leaving 189 populations in the primary set, including 161 with complete 11-gene profiles. Of these, 173 populations had at least 50 individuals, including 146 with complete profiles.

**Supplementary Figure 2**. Pairwise regional differentiation

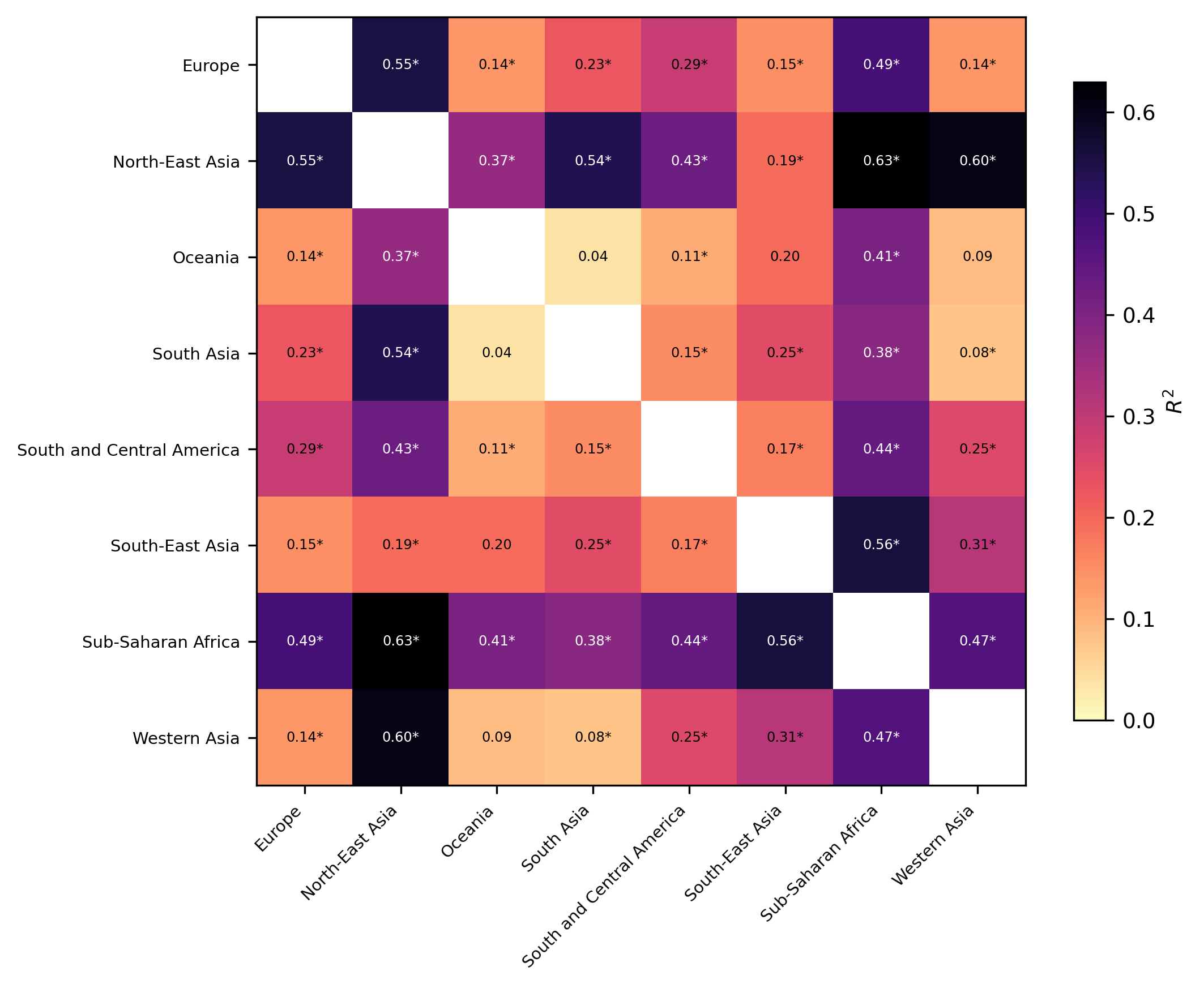

**Caption:** Proportion of variation explained in pairwise comparisons between geographic regions represented by at least three complete profiles, estimated using distance-based permutation analyses. Asterisks indicate comparisons that remained significant after Benjamini–Hochberg false-discovery-rate correction. **Abbreviations:** R²: proportion of variation explained; FDR: false discovery rate.

**Supplementary Figure 3.** Selection of the number of clusters
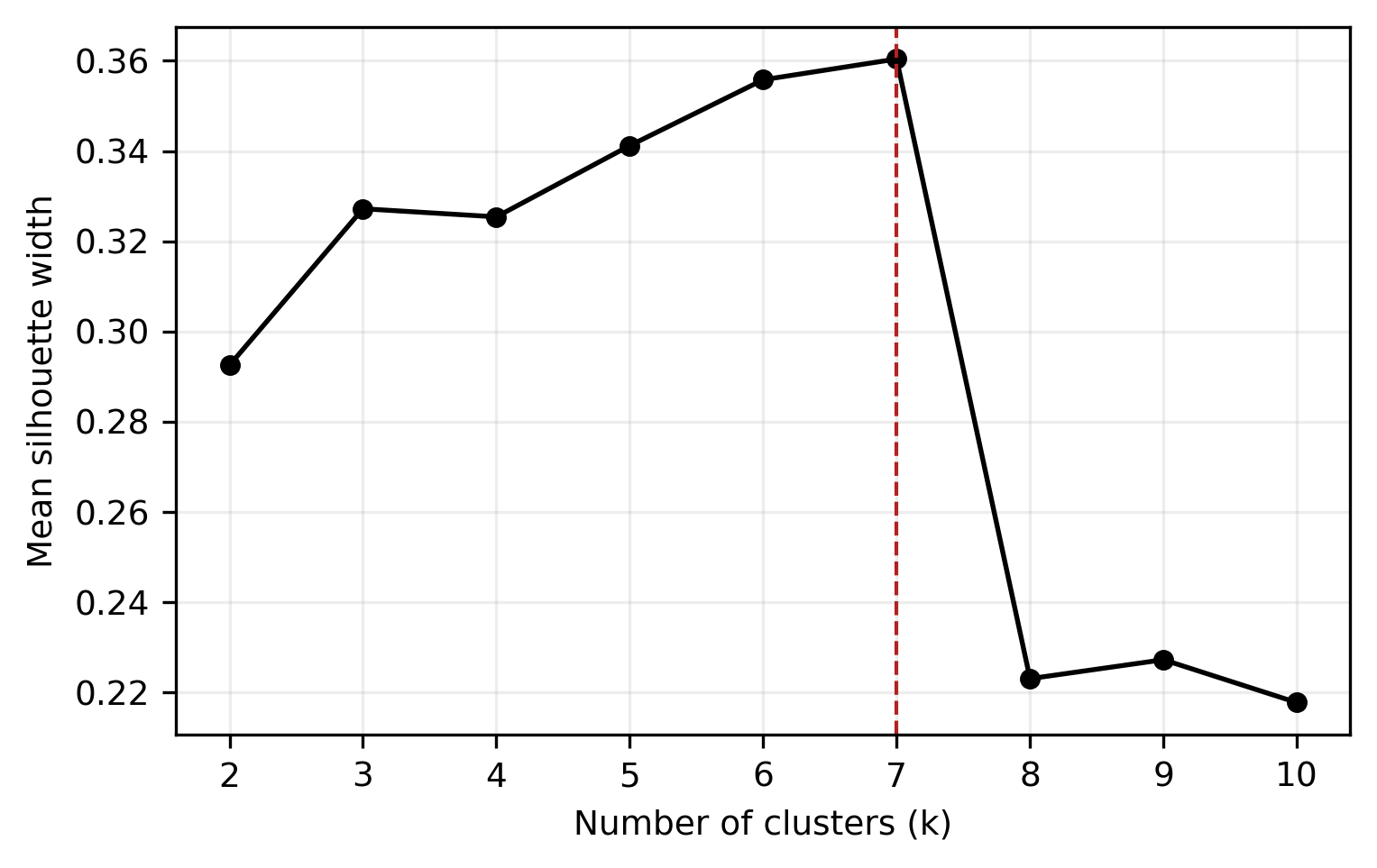
**Caption:** Mean silhouette width across hierarchical clustering solutions containing 2–10 clusters. The dashed vertical line indicates the selected seven-cluster solution. Exact values are provided in Supplementary Table 6. **Abbreviations:** k: number of clusters.

**Supplementary Figure 4.** Gene-specific differentiation.
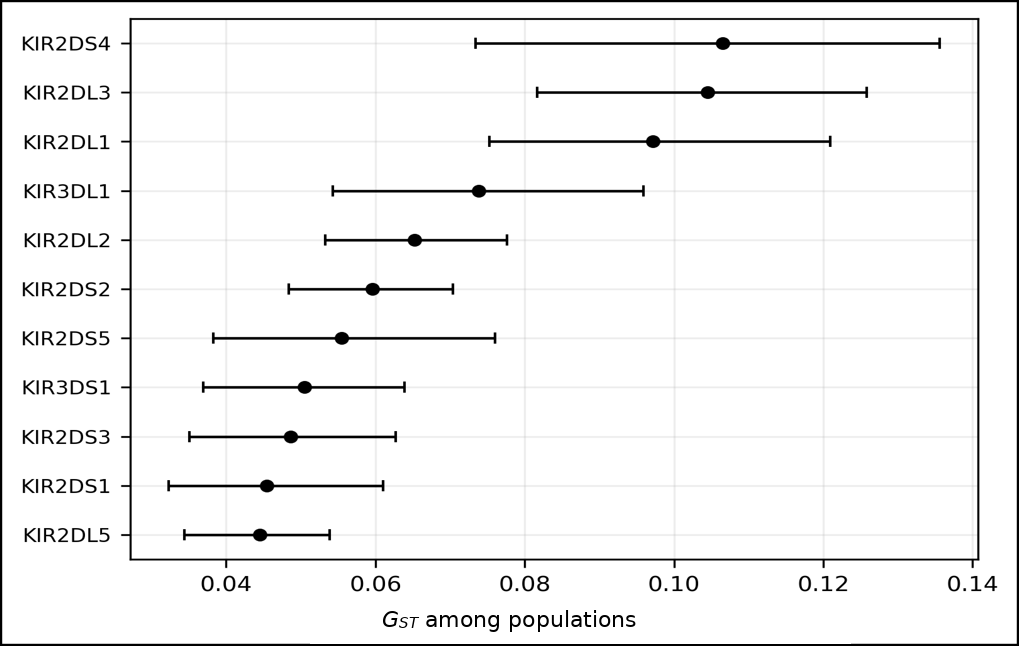
**Caption:** Differentiation among the 161 complete-profile populations for each of the 11 variable KIR genes. Points show Nei’s coefficient of gene differentiation, and horizontal lines show percentile 95% confidence intervals obtained from 1,000 bootstrap samples generated by resampling populations with replacement. Exact estimates are provided in Supplementary Table 7. **Abbreviations**: G_ST_: coefficient of gene differentiation.

**Supplementary Figure 5.** Expected heterozygosity and distance from East Africa

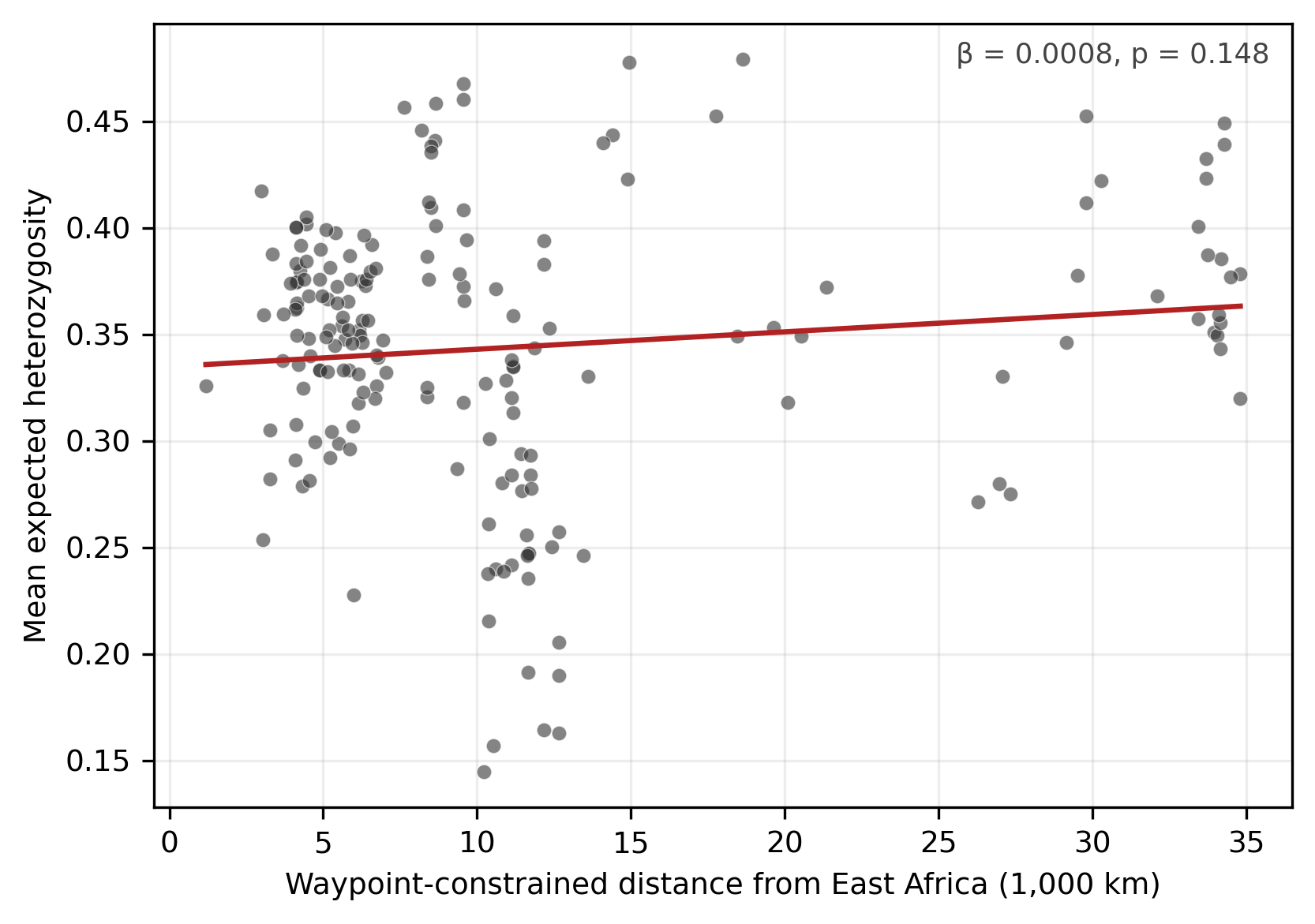

**Caption:** Mean expected heterozygosity plotted against waypoint-constrained distance from East Africa in the primary analysis set. The fitted line is unadjusted, whereas the reported coefficient is from the model adjusted for log-transformed sample size. Exact estimates for all four distance definitions are provided in Supplementary Table 8. **Abbreviations:** β: regression coefficient; He: expected heterozygosity.

**Supplementary Figure 6.** Within-region latitude coefficients

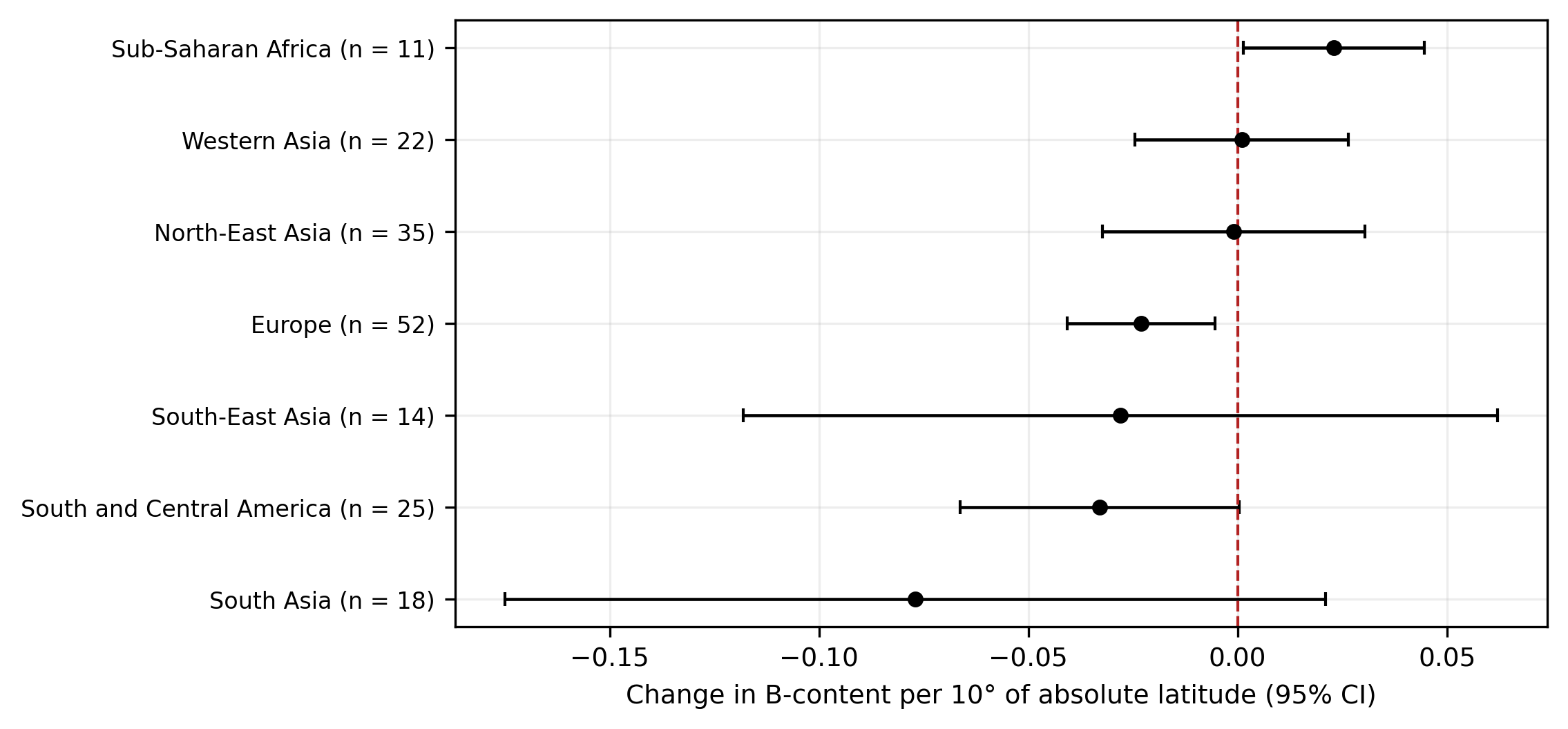

**Caption:** Coefficients and 95% confidence intervals for the association between B-content and absolute latitude, expressed per 10° increase in latitude and estimated separately within each geographic region represented by at least 10 populations. No association remained significant after false-discovery-rate correction. Exact estimates are provided in Supplementary Table 11. **Abbreviations:** n: number of populations; CI: confidence interval.

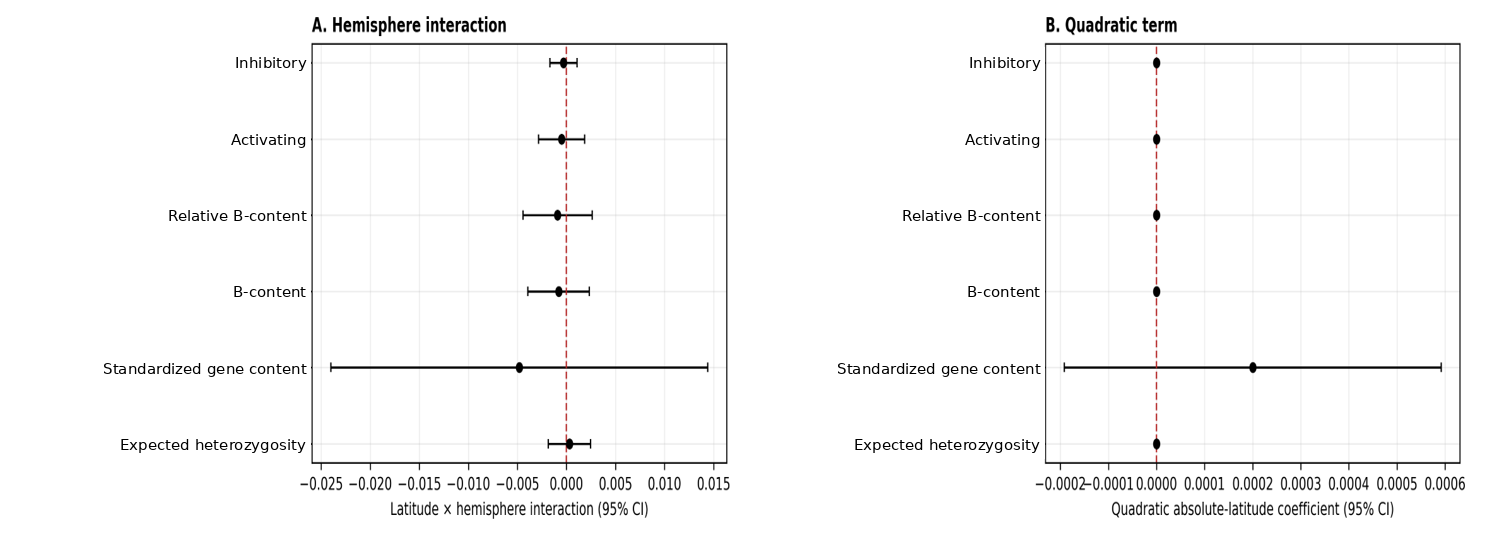
**Supplementary Figure 7.** Secondary latitude specifications

**Caption:** (A) Coefficients and 95% confidence intervals for the interaction between absolute latitude and hemisphere across the gene-content and diversity measures. (B) Coefficients and 95% confidence intervals for the quadratic absolute-latitude term from models also containing the linear absolute-latitude term and log-transformed sample size. Dashed vertical lines indicate no association. Uneven hemispheric coverage limited estimation of hemisphere-specific slopes. Exact estimates are provided in Supplementary Table 16. **Abbreviations:** β: regression coefficient; CI: confidence interval.

**Supplementary Figure 8.** Residual spatial autocorrelation.

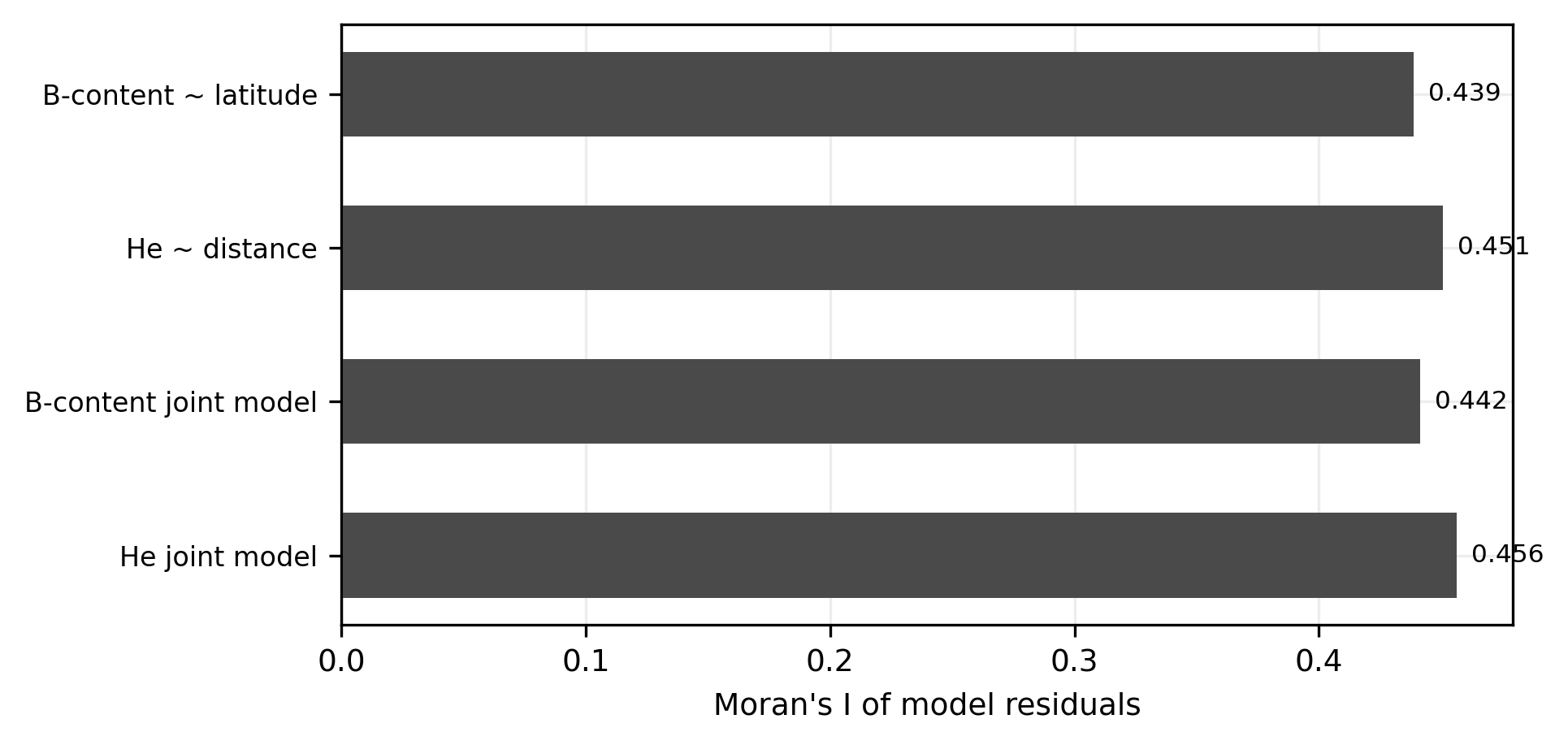

**Caption:** Moran’s I for residuals from the primary latitude, distance, and joint latitude–distance models, calculated using a row-standardized eight-nearest-neighbour spatial network. All permutation p-values were 0.0010. Exact estimates are provided in Supplementary Table 17. **Abbreviations:** I: Moran’s I; He: expected heterozygosity.

**Supplementary Figure 9.** Robustness and power analyses results.

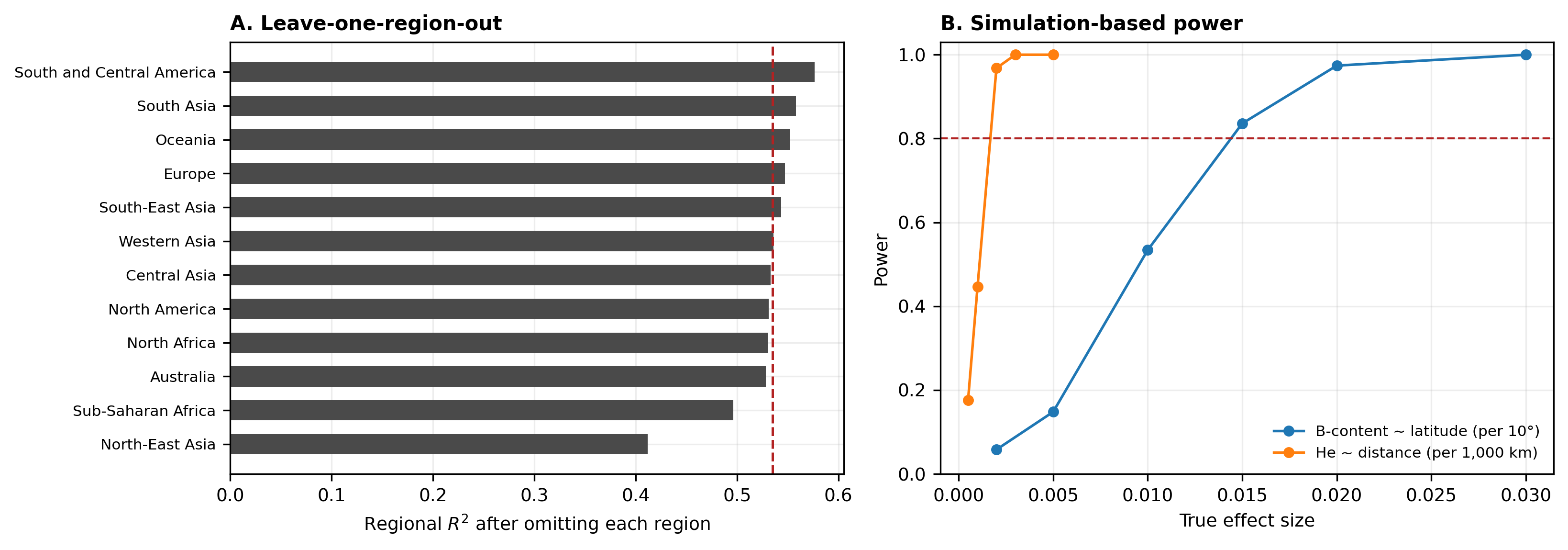

**Caption:** (A) Proportion of multivariate variation attributable to geographic region after sequentially omitting each region. The dashed vertical line indicates the full-data estimate (R²=0.5349). (B) Simulation-based power curves for the two primary regression models: B-content on absolute latitude per 10° and expected heterozygosity on waypoint-constrained distance per 1,000 km. The dashed horizontal line indicates 80% power. Exact results are provided in Supplementary Table 19 for panel A and Supplementary Table 13 for panel B. **Abbreviations:** R²: proportion of variation explained; He: expected heterozygosity.

**Supplementary Table 1.** populations from National Allele Frequency Database included in the study.

| Population | Abbreviation | Region | Broad area | Coordinates (°N, °E) | Sample size | Genes reported | Analysis set | Source publication |
| --- | --- | --- | --- | --- | --- | --- | --- | --- |
| Australia Queensland Brisbane | AUS-Bris | Australia | Oceania | -27.467, 153.017 | 106 | 11 | Primary analysis | Seymour 2014 |
| Australia South Aborigine | AUS-Aborig | Australia | Oceania | -33.917, 151.200 | 67 | 9 | Primary analysis | Toneva 2001 |
| Uzbekistan | UZB | Central Asia | Asia | 41.267, 69.217 | 67 | 11 | Primary analysis | Evseeva |
| Belgium | BEL-1 | Europe | Europe | 50.000, 5.000 | 148 | 10 | Primary analysis | Demanet 2004 |
| Belgium pop 2 | BEL-2 | Europe | Europe | 51.033, 3.050 | 100 | 10 | Primary analysis | Hidajat 2004 |
| Bulgaria | BGR | Europe | Europe | 42.700, 23.333 | 54 | 10 | Primary analysis | Naumova |
| Croatia | HRV-1 | Europe | Europe | 45.800, 15.967 | 125 | 11 | Primary analysis | Burek Kamenaric 2013 |
| Croatia pop 2 | HRV-2 | Europe | Europe | 45.800, 15.967 | 111 | 11 | Primary analysis | Burek Kamenaric |
| Croatia pop 3 | HRV-3 | Europe | Europe | 45.800, 15.950 | 121 | 11 | Primary analysis | Burek 2013 |
| Czech Republic | CZE | Europe | Europe | 50.083, 14.417 | 125 | 11 | Primary analysis | Pavlova 2008 |
| England | ENG-1 | Europe | Europe | 51.000, 0.000 | 136 | 10 | Primary analysis | Norman 2007 |
| England pop 3 | ENG-3 | Europe | Europe | 51.750, -1.250 | 334 | 11 | Primary analysis | Harvey |
| England pop 4 | ENG-4 | Europe | Europe | 51.500, -0.117 | 99 | 11 | Primary analysis | Navarette |
| England pop 5 | ENG-5 | Europe | Europe | 50.817, -1.083 | 584 | 11 | Primary analysis | Hiby 2010 |
| England Oxfordshire | ENG-Oxf | Europe | Europe | 51.750, -1.250 | 216 | 11 | Primary analysis | Jones 2006 |
| England West Midlands | ENG-WMid | Europe | Europe | 52.917, -1.900 | 126 | 11 | Primary analysis | Cook 2003 |
| Finland Helsinki | FIN-Hels | Europe | Europe | 60.000, 25.000 | 101 | 11 | Primary analysis | Denis 2005 |
| France Reims | FRA-Reims | Europe | Europe | 49.250, 4.033 | 102 | 11 | Primary analysis | Levinson 2008 |
| France Southeast | FRA-SE1 | Europe | Europe | 46.000, 5.000 | 130 | 9 | Primary analysis | Dubois 2004 |
| France Southeast pop 2 | FRA-SE2 | Europe | Europe | 43.283, 5.367 | 38 | 11 | Primary analysis | Frassati 2006 |
| France West | FRA-West | Europe | Europe | 47.000, 2.000 | 108 | 11 | Primary analysis | Denis 2005 |
| Germany Hesse | DEU-Hesse | Europe | Europe | 50.650, 8.583 | 99 | 10 | Primary analysis | Becker 2003 |
| Germany | DEU-1 | Europe | Europe | 52.367, 9.733 | 100 | 9 | Primary analysis | Momot 2004 |
| Greece | GRC | Europe | Europe | 37.967, 23.717 | 233 | 10 | Primary analysis | Niokou 2003 |
| Ireland pop 2 | IRL | Europe | Europe | 52.433, -8.433 | 240 | 11 | Primary analysis | Keating 2015 |
| Ireland Northern | NIR-1 | Europe | Europe | 54.667, -6.750 | 200 | 11 | Primary analysis | AFND only |
| Ireland Northern pop2 | NIR-2 | Europe | Europe | 54.667, -6.750 | 154 | 11 | Primary analysis | AFND only |
| Italy | ITA-1 | Europe | Europe | 44.500, 11.350 | 217 | 9 | Primary analysis | Bontadini 2005 |
| Italy Lombardy | ITA-Lomb | Europe | Europe | 45.000, 9.000 | 50 | 11 | Primary analysis | Martinetti |
| Italy Sardinia Cagliari | ITA-Sard | Europe | Europe | 39.217, 9.100 | 181 | 11 | Primary analysis | Littera 2013 |
| Italy West Sicily | ITA-Sicily | Europe | Europe | 38.000, 13.300 | 60 | 9 | Primary analysis | Gambino 2018 |
| Latvia | LVA | Europe | Europe | 57.000, 25.000 | 100 | 11 | Primary analysis | Nikitina-Zake 2004 |
| Macedonia | MKD | Europe | Europe | 42.000, 21.000 | 214 | 11 | Primary analysis | Spiroski 2010 |
| Macedonia Romani | MKD-Romani | Europe | Europe | 41.983, 21.417 | 103 | 11 | Primary analysis | Petlichkovski 2011 |
| Norway | NOR | Europe | Europe | 59.933, 10.750 | 368 | 11 | Primary analysis | Karlsen 2007 |
| Poland | POL-1 | Europe | Europe | 53.417, 14.583 | 690 | 11 | Primary analysis | Majorczyk 2007 |
| Poland pop2 | POL-2 | Europe | Europe | 51.117, 17.033 | 60 | 11 | Primary analysis | Karabon 2011 |
| Portugal Azores Terceira Island | PRT-Azores | Europe | Europe | 38.650, -27.217 | 127 | 11 | Primary analysis | Bruges-Armas |
| Portugal | PRT | Europe | Europe | 38.767, -9.183 | 65 | 11 | Primary analysis | Santos |
| Romania | ROU | Europe | Europe | 44.417, 26.100 | 123 | 10 | Primary analysis | Constantinescu 2006 |
| Russia Chechens | RUS-Chech | Europe | Europe | 45.000, 46.000 | 48 | 11 | Primary analysis | Khamaganova (conference abstract) |
| Russia Moscow | RUS-Mosc | Europe | Europe | 55.650, 37.483 | 84 | 11 | Primary analysis | Zakharova 2017 |
| Russia Northwest pop 3 | RUS-NW | Europe | Europe | 59.967, 30.300 | 100 | 11 | Primary analysis | Bubnova 2012 (conference abstract) |
| Scotland Orkney | SCO-Orkney | Europe | Europe | 59.000, -3.000 | 90 | 11 | Primary analysis | Tonks |
| Serbia Vojvodina | SRB-Vojv | Europe | Europe | 45.250, 19.850 | 134 | 11 | Primary analysis | AFND only |
| Slovenia | SVN | Europe | Europe | 46.250, 15.017 | 130 | 10 | Primary analysis | Smigoc Schweiger |
| Spain Avilés | ESP-Aviles | Europe | Europe | 43.150, -5.917 | 339 | 11 | Primary analysis | Diaz-Pena 2016 |
| Spain Basque | ESP-Basque | Europe | Europe | 43.250, -2.917 | 71 | 11 | Primary analysis | Santin 2006 |
| Spain Cantabria | ESP-Cantab | Europe | Europe | 43.467, -3.800 | 230 | 11 | Primary analysis | Ocejo-Vinals |
| Spain Granada | ESP-Granada | Europe | Europe | 37.167, -3.600 | 100 | 11 | Primary analysis | Middleton |
| Spain Madrid | ESP-Madrid | Europe | Europe | 40.400, -3.683 | 131 | 9 | Primary analysis | Vilches |
| Spain Murcia | ESP-Murcia | Europe | Europe | 38.000, -1.100 | 200 | 11 | Primary analysis | Campillo 2013 |
| Spain Sevilla | ESP-Sevilla | Europe | Europe | 37.367, -5.967 | 278 | 11 | Primary analysis | AFND only |
| Sweden | SWE | Europe | Europe | 59.317, 18.067 | 102 | 11 | Primary analysis | AFND only |
| Sweden Vasterbotten | SWE-Vaster | Europe | Europe | 59.350, 18.067 | 150 | 11 | Primary analysis | Arnheim 2005 |
| Morocco Chaouya | MAR-Chaouya | North Africa | Africa | 33.067, -7.617 | 67 | 11 | Primary analysis | Canossi |
| Tunisia | TUN | North Africa | Africa | 34.117, 9.550 | 114 | 10 | Primary analysis | AFND only |
| USA Eastern | USA-East | North America | North America | 39.000, -76.700 | 213 | 11 | Primary analysis | Carrington 2005 |
| China Eastern Mainland Han | CHN-Han-E | North-East Asia | Asia | 29.000, 120.000 | 106 | 11 | Primary analysis | Wu 2009 |
| China Guangdong Han | CHN-Han-Gd | North-East Asia | Asia | 23.933, 114.883 | 107 | 11 | Primary analysis | Cai 2012 |
| China Hubei Province Tujia | CHN-Tujia | North-East Asia | Asia | 30.283, 109.467 | 124 | 11 | Primary analysis | Wang 2016 |
| China Hunan Han | CHN-Han-Hn | North-East Asia | Asia | 28.117, 111.433 | 448 | 11 | Primary analysis | Cai 2012 |
| China Inner Mongolia Han | CHN-Han-IM | North-East Asia | Asia | 42.883, 113.567 | 104 | 11 | Primary analysis | Cai 2012 |
| China Inner Mongolia Mongol | CHN-Mongol | North-East Asia | Asia | 42.883, 113.567 | 160 | 11 | Primary analysis | Cai 2012 |
| China Inner Mongolia Province pop 2 | CHN-IMong2 | North-East Asia | Asia | 44.000, 113.000 | 87 | 11 | Primary analysis | Wang 2012 |
| China Jiangsu Province Han | CHN-Han-Js1 | North-East Asia | Asia | 31.300, 120.600 | 150 | 11 | Primary analysis | He 2010 |
| China Jiangsu Province Han pop2 | CHN-Han-Js2 | North-East Asia | Asia | 33.000, 120.000 | 60 | 11 | Primary analysis | AFND only |
| China Mongolia Province | CHN-MongP | North-East Asia | Asia | 43.617, 122.250 | 90 | 11 | Primary analysis | Jiang 2013 |
| China Nanjing | CHN-Nanj | North-East Asia | Asia | 32.050, 118.767 | 238 | 11 | Primary analysis | Pan |
| China Shaanxi Province Han | CHN-Han-Sx | North-East Asia | Asia | 35.600, 108.400 | 104 | 11 | Primary analysis | Zhu 2011 |
| China Shenzhen Han pop 2 | CHN-Han-Sz | North-East Asia | Asia | 22.550, 114.050 | 503 | 11 | Primary analysis | Zhen 2014 |
| China Tibet Lhasa | CHN-Tibet | North-East Asia | Asia | 29.650, 91.100 | 102 | 10 | Primary analysis | Wang 2010 |
| China Xinjiang Region Urumqi Uygur | CHN-Uygur | North-East Asia | Asia | 43.817, 87.600 | 120 | 11 | Primary analysis | Zhu 2012 |
| China Yunnan Han | CHN-Han-Yn | North-East Asia | Asia | 25.033, 102.700 | 404 | 11 | Primary analysis | Shi 2013 |
| China Yunnan Province Dali Bai | CHN-Bai | North-East Asia | Asia | 25.600, 100.267 | 100 | 11 | Primary analysis | Zhu 2011 |
| China Yunnan Province Drung | CHN-Drung | North-East Asia | Asia | 25.050, 101.867 | 152 | 11 | Primary analysis | Jiang 2017 |
| China Yunnan Province Fengyandong Han | CHN-Han-Fy | North-East Asia | Asia | 24.050, 105.067 | 93 | 11 | Primary analysis | Shi 2011 |
| China Yunnan Province Fugong Nu | CHN-Nu | North-East Asia | Asia | 25.900, 98.867 | 112 | 11 | Primary analysis | Shi 2011 |
| China Yunnan Province Menghai Bulang | CHN-Bulang | North-East Asia | Asia | 21.967, 100.467 | 109 | 11 | Primary analysis | Shi 2011 |
| China Yunnan Province Sunan Yugu | CHN-Yugu | North-East Asia | Asia | 38.833, 99.617 | 104 | 11 | Primary analysis | Shi 2011 |
| China Yunnan Province Tiandeng Zhuang | CHN-Zhuang | North-East Asia | Asia | 22.850, 107.383 | 98 | 11 | Primary analysis | Shi 2011 |
| China Zhejiang Han | CHN-Han-Zj | North-East Asia | Asia | 29.200, 120.500 | 273 | 11 | Primary analysis | Tao 2017 |
| China Zhejiang Province Han | CHN-Han-Zj2 | North-East Asia | Asia | 30.250, 120.167 | 104 | 10 | Primary analysis | Jiang 2005 |
| Hong Kong Chinese | HKG | North-East Asia | Asia | 22.000, 114.000 | 100 | 11 | Primary analysis | Williams 2004 |
| Japan | JPN-1 | North-East Asia | Asia | 35.683, 139.767 | 50 | 11 | Primary analysis | Suzuki 2004 |
| Japan pop 2 | JPN-2 | North-East Asia | Asia | 35.683, 139.767 | 41 | 11 | Primary analysis | Yawata 2002 |
| Japan pop 3 | JPN-3 | North-East Asia | Asia | 35.683, 139.767 | 132 | 11 | Primary analysis | Yawata 2006 |
| Japan Kyoto | JPN-Kyoto | North-East Asia | Asia | 35.017, 135.767 | 240 | 11 | Primary analysis | Mogami 2007 |
| Japan Tokyo | JPN-Tokyo | North-East Asia | Asia | 35.683, 139.767 | 239 | 11 | Primary analysis | Miyashita 2006 |
| Korea pop 2 | KOR-2 | North-East Asia | Asia | 37.550, 126.950 | 159 | 11 | Primary analysis | Kim 2014 |
| South Korea | KOR-1 | North-East Asia | Asia | 37.000, 126.000 | 154 | 11 | Primary analysis | Whang 2005 |
| Taiwan Han pop 4 | TWN-Han | North-East Asia | Asia | 23.767, 121.000 | 97 | 11 | Primary analysis | Wu 2009 |
| Taiwan | TWN-1 | North-East Asia | Asia | 23.767, 121.000 | 96 | 9 | Primary analysis | Yen 2006 |
| Cook Islands | COK | Oceania | Oceania | -21.200, -159.767 | 48 | 11 | Primary analysis | Velickovic 2006 |
| Papua New Guinea Irian Jaya | PNG-IrianJ | Oceania | Oceania | -1.000, 133.000 | 50 | 11 | Primary analysis | Velickovic 2009 |
| Samoa | WSM | Oceania | Oceania | -13.833, -171.750 | 50 | 11 | Primary analysis | Velickovic 2006 |
| Solomon Islands | SLB | Oceania | Oceania | -9.467, 159.817 | 40 | 11 | Primary analysis | Taniguchi 2009 |
| Tokelau | TKL | Oceania | Oceania | -9.167, -171.833 | 47 | 11 | Primary analysis | Velickovic 2006 |
| Tonga | TON | Oceania | Oceania | -21.133, -175.200 | 49 | 11 | Primary analysis | Velickovic 2006 |
| India Madurai Piramalai Kallar Dravidian | IND-PKallar | South Asia | Asia | 9.100, 78.017 | 100 | 11 | Primary analysis | Maruthamuthu 2015 |
| India Maharashtra Pune Western | IND-Pune | South Asia | Asia | 18.517, 73.850 | 104 | 10 | Primary analysis | Alagarasu 2015 |
| India Mumbai Maharashtrian | IND-Mahar | South Asia | Asia | 18.967, 72.817 | 139 | 11 | Primary analysis | Kulkarni 2008 |
| India Mumbai Parsi | IND-Parsi | South Asia | Asia | 18.967, 72.817 | 145 | 11 | Primary analysis | Kulkarni 2008 |
| India North Hindu | IND-N-Hindu | South Asia | Asia | 25.000, 74.000 | 72 | 11 | Primary analysis | Rajalingam 2002 |
| India North | IND-North1 | South Asia | Asia | 26.867, 80.967 | 512 | 11 | Primary analysis | Agrawal 2013 |
| India North pop 2 | IND-North2 | South Asia | Asia | 26.850, 80.917 | 100 | 11 | Primary analysis | Prakash 2014 |
| India Northeast Assam Adivasi | IND-Adivasi | South Asia | Asia | 21.133, 72.917 | 101 | 11 | Primary analysis | Dutta 2013 |
| India Northeast Assam Ahom | IND-Ahom | South Asia | Asia | 21.133, 72.917 | 104 | 11 | Primary analysis | Dutta 2013 |
| India Northeast Assam | IND-Assam | South Asia | Asia | 26.483, 91.667 | 177 | 11 | Primary analysis | Dutta 2014 |
| India Northeast Assam Kharachi | IND-Kharachi | South Asia | Asia | 21.133, 72.917 | 108 | 11 | Primary analysis | Dutta 2013 |
| India Tamil Nadu Kanikar | IND-Kanikar | South Asia | Asia | 13.150, 80.450 | 35 | 11 | Primary analysis | Rajalingam 2008 |
| India Tamil Nadu Mollukurumba | IND-Mollu | South Asia | Asia | 13.150, 80.450 | 41 | 11 | Primary analysis | Rajalingam 2008 |
| India Tamil Nadu Paravar | IND-Paravar | South Asia | Asia | 13.150, 80.450 | 77 | 11 | Primary analysis | Rajalingam 2008 |
| India West Bengal Rabha | IND-Rabha | South Asia | Asia | 26.317, 89.667 | 50 | 11 | Primary analysis | Guha 2015 |
| India West Bengal Rajbanshi | IND-Rajban | South Asia | Asia | 22.550, 88.367 | 75 | 11 | Primary analysis | Guha 2013 |
| India Western | IND-West | South Asia | Asia | 19.000, 72.833 | 161 | 11 | Primary analysis | D Silva 2019 |
| Pakistan Karachi | PAK-Karachi | South Asia | Asia | 24.850, 67.000 | 78 | 11 | Primary analysis | Norman 2002 |
| Argentina Buenos Aires | ARG-BsAs | South and Central America | South America | -34.633, -58.450 | 365 | 11 | Primary analysis | Marcos |
| Argentina Chaco Region Wichi | ARG-Wichi-Ch | South and Central America | South America | -24.000, -61.000 | 82 | 11 | Primary analysis | Marcos |
| Argentina Chiriguano | ARG-Chirig | South and Central America | South America | -23.133, -64.050 | 54 | 11 | Primary analysis | Flores |
| Argentina | ARG | South and Central America | South America | -34.600, -58.367 | 102 | 11 | Primary analysis | AFND only |
| Argentina Salta Wichi | ARG-Wichi-Sa | South and Central America | South America | -23.133, -64.033 | 19 | 11 | Primary analysis | Flores |
| Brazil Amazon | BRA-Amaz | South and Central America | South America | -6.817, -35.750 | 40 | 11 | Primary analysis | Ewerton 2007 |
| Brazil Bahia Ilhéus | BRA-Ilheus | South and Central America | South America | -14.783, -39.033 | 102 | 10 | Primary analysis | Issler 2018 |
| Brazil Curitiba | BRA-Curit | South and Central America | South America | -25.417, -49.267 | 164 | 11 | Primary analysis | Augusto 2011 |
| Brazil Guarani Kaiowá | BRA-Gua-Kai | South and Central America | South America | -23.100, -55.200 | 96 | 11 | Primary analysis | Augusto 2013 |
| Brazil Guarani MByá | BRA-Gua-MBy | South and Central America | South America | -25.300, -52.533 | 81 | 11 | Primary analysis | Augusto 2013 |
| Brazil Guarani Ñandeva | BRA-Gua-Nan | South and Central America | South America | -23.800, -54.500 | 50 | 11 | Primary analysis | Augusto 2013 |
| Brazil Kaingang | BRA-Kaing | South and Central America | South America | -25.300, -52.533 | 100 | 11 | Primary analysis | Augusto 2013 |
| Brazil Mato Grosso do Sul Campo Grande | BRA-CGrande | South and Central America | South America | -20.450, -54.600 | 206 | 11 | Primary analysis | Augusto 2012 |
| Brazil Paraná Maringa | BRA-Maring | South and Central America | South America | -23.417, -51.933 | 130 | 11 | Primary analysis | Caniatti 2018 |
| Brazil Rondonia Porto Velho | BRA-PVelho | South and Central America | South America | -8.750, -63.900 | 377 | 11 | Primary analysis | Perce-da-Silva 2015 |
| Brazil Sao Paulo | BRA-SPaulo | South and Central America | South America | -20.967, -49.117 | 289 | 11 | Primary analysis | Marangon 2011 |
| Chile Talca | CHL-Talca | South and Central America | South America | -34.867, -71.667 | 90 | 11 | Primary analysis | Diaz-Pena 2018 |
| Costa Rica Guanacaste | CRI-Guanac | South and Central America | Central America | 9.933, -84.083 | 117 | 11 | Primary analysis | Carrington 2005 |
| Mexico Huichol | MEX-Huichol | South and Central America | Central America | 21.733, -105.217 | 73 | 11 | Primary analysis | Gutierrez-Rodriguez 2006 |
| Mexico Purepecha | MEX-Purep | South and Central America | Central America | 19.767, -101.183 | 53 | 11 | Primary analysis | Gutierrez-Rodriguez 2006 |
| Mexico Tarahumara | MEX-Tarah | South and Central America | Central America | 27.500, -107.750 | 65 | 11 | Primary analysis | Gutierrez-Rodriguez 2006 |
| Panama | PAN | South and Central America | Central America | 8.967, -79.533 | 116 | 11 | Primary analysis | Vernaza 2013 (conference abstract) |
| Venezuela Bari | VEN-Bari | South and Central America | South America | 10.000, -72.950 | 80 | 11 | Primary analysis | Gendzekhadze 2006 |
| Venezuela Warao | VEN-Warao | South and Central America | South America | 8.500, -67.167 | 89 | 11 | Primary analysis | Gendzekhadze 2006 |
| Venezuela Yucpa | VEN-Yucpa | South and Central America | South America | 10.000, -72.950 | 61 | 11 | Primary analysis | Gendzekhadze 2006 |
| Borneo Kalimantan | IDN-Kalim | South-East Asia | Asia | -2.217, 113.917 | 48 | 11 | Primary analysis | AFND only |
| East Timor | TLS | South-East Asia | Asia | -8.567, 125.567 | 50 | 11 | Primary analysis | Velickovic 2008 |
| Indonesia Java | IDN-Java | South-East Asia | Asia | -7.500, 111.250 | 45 | 11 | Primary analysis | Velickovic 2009 |
| Indonesia Moluccan Islands | IDN-Moluc | South-East Asia | Asia | 0.500, 127.583 | 63 | 10 | Primary analysis | Velickovic 2007 |
| Indonesia Sulawesi Minahasans | IDN-Minah | South-East Asia | Asia | -4.167, 120.000 | 60 | 10 | Primary analysis | Velickovic 2007 |
| Indonesia West Sumatra Minangs | IDN-Minang | South-East Asia | Asia | -0.600, 99.850 | 45 | 9 | Primary analysis | Velickovic 2007 |
| Malaysia Peninsular | MYS-Pen | South-East Asia | Asia | 6.083, 102.283 | 120 | 11 | Primary analysis | NurWaliyuddin 2014 |
| Singapore Malays | SGP-Malay | South-East Asia | Asia | 1.367, 103.800 | 80 | 11 | Primary analysis | Lee 2008 |
| Thailand Bangkok | THA-Bkk1 | South-East Asia | Asia | 13.750, 100.483 | 119 | 10 | Primary analysis | Norman 2007 |
| Thailand Bangkok pop 2 | THA-Bkk2 | South-East Asia | Asia | 13.750, 100.483 | 100 | 11 | Primary analysis | Vejbaesya |
| Thailand Bangkok pop 3 | THA-Bkk3 | South-East Asia | Asia | 13.750, 100.517 | 150 | 11 | Primary analysis | Vejbaesya 2014 |
| Thailand North East | THA-NE | South-East Asia | Asia | 16.467, 102.817 | 235 | 11 | Primary analysis | Chaisri 2013 |
| Vietnam Hanoi | VNM-Hanoi1 | South-East Asia | Asia | 21.033, 105.850 | 59 | 9 | Primary analysis | Toneva 2001 |
| Vietnam Hanoi pop 2 | VNM-Hanoi | South-East Asia | Asia | 21.017, 105.750 | 140 | 11 | Primary analysis | Amorim 2018 |
| Equatorial Guinea Bioko Island Bubi | GNQ-Bubi | Sub-Saharan Africa | Africa | 1.917, 10.100 | 95 | 11 | Primary analysis | AFND only |
| Gabon | GAB | Sub-Saharan Africa | Africa | -1.633, 13.583 | 54 | 11 | Primary analysis | Wauquier 2010 |
| Ghana | GHA | Sub-Saharan Africa | Africa | 5.550, -0.250 | 41 | 11 | Primary analysis | Navarette |
| Ivory Coast Abidjan | CIV-Abidj | Sub-Saharan Africa | Africa | 5.333, -4.017 | 25 | 11 | Primary analysis | Jennes 2006 |
| Senegal | SEN | Sub-Saharan Africa | Africa | 14.000, -16.000 | 118 | 11 | Primary analysis | Denis 2005 |
| South Africa Black African | ZAF-Black | Sub-Saharan Africa | Africa | -26.133, 28.050 | 167 | 11 | Primary analysis | Gentle 2017 |
| South Africa Carletonville Gauteng | ZAF-Carlet | Sub-Saharan Africa | Africa | -26.350, 27.383 | 81 | 11 | Primary analysis | Hong 2011 |
| South Africa San | ZAF-San | Sub-Saharan Africa | Africa | -34.350, 18.467 | 91 | 11 | Primary analysis | Middleton |
| South Africa Xhosa | ZAF-Xhosa | Sub-Saharan Africa | Africa | -34.000, 19.000 | 50 | 11 | Primary analysis | Williams 2004 |
| Uganda Kampala | UGA-Kamp | Sub-Saharan Africa | Africa | 0.300, 32.567 | 492 | 11 | Primary analysis | Nakimuli 2013 |
| Zimbabwe Harare | ZWE-Harare | Sub-Saharan Africa | Africa | -19.133, 29.883 | 97 | 11 | Primary analysis | Mhandire 2016 |
| Georgia | GEO | Western Asia | Middle East | 41.717, 44.783 | 188 | 11 | Primary analysis | Norman 2007 |
| Iran East Azerbaijan Azerbaijani | IRN-Azerb | Western Asia | Middle East | 38.067, 46.283 | 100 | 11 | Primary analysis | Hiby 2010 |
| Iran East Azerbaijan Azeri | IRN-Azeri | Western Asia | Middle East | 38.067, 46.300 | 84 | 11 | Primary analysis | Ashouri 2009 |
| Iran Fars Persian | IRN-Persian | Western Asia | Middle East | 29.600, 52.517 | 248 | 11 | Primary analysis | Ashouri 2009 |
| Iran Hormozgan and Balochistan Jonobi | IRN-Jonobi | Western Asia | Middle East | 27.183, 56.267 | 100 | 11 | Primary analysis | Hiby 2010 |
| Iran | IRN | Western Asia | Middle East | 35.683, 51.417 | 200 | 11 | Primary analysis | Tajik |
| Iran Khuzestan Arab | IRN-Arab | Western Asia | Middle East | 31.300, 48.700 | 76 | 11 | Primary analysis | Ashouri 2009 |
| Iran Khuzestan Bakhtiari | IRN-Bakht | Western Asia | Middle East | 31.300, 48.700 | 96 | 11 | Primary analysis | Ashouri 2009 |
| Iran Lur | IRN-Lur | Western Asia | Middle East | 33.500, 48.667 | 100 | 11 | Primary analysis | Shahsavar 2013 |
| Iran Northern | IRN-North | Western Asia | Middle East | 35.683, 51.417 | 100 | 11 | Primary analysis | Amirzargar |
| Iran Shiraz | IRN-Shiraz | Western Asia | Middle East | 29.583, 52.567 | 273 | 10 | Primary analysis | Barani 2018 |
| Iran Tehran | IRN-Tehr | Western Asia | Middle East | 35.700, 51.417 | 100 | 11 | Primary analysis | Tajik 2012 |
| Jordan Palestine | JOR-Pal | Western Asia | Middle East | 31.583, 36.333 | 105 | 11 | Primary analysis | Norman 2001 |
| Lebanon | LBN | Western Asia | Middle East | 33.900, 35.533 | 120 | 11 | Primary analysis | Mahfouz 2006 |
| Oman | OMN | Western Asia | Middle East | 22.000, 56.000 | 99 | 11 | Primary analysis | Williams 2004 |
| Saudi Arabia | SAU-1 | Western Asia | Middle East | 24.700, 46.717 | 162 | 11 | Primary analysis | Sheereen |
| Saudi Arabia pop 2 | SAU-2 | Western Asia | Middle East | 24.717, 46.800 | 148 | 11 | Primary analysis | Osman 2014 |
| Saudi Arabia pop 3 | SAU-3 | Western Asia | Middle East | 25.000, 45.000 | 65 | 10 | Primary analysis | Alomar 2017 |
| Saudi Arabia Riyadh | SAU-Riyadh | Western Asia | Middle East | 24.633, 46.717 | 152 | 11 | Primary analysis | Osman 2017 |
| Turkey pop 2 | TUR-2 | Western Asia | Middle East | 41.017, 28.983 | 154 | 11 | Primary analysis | Middleton 2007 |
| Turkey pop 3 | TUR-3 | Western Asia | Middle East | 40.900, 29.083 | 187 | 10 | Primary analysis | AFND only |
| Turkey South | TUR-South | Western Asia | Middle East | 37.350, 34.600 | 200 | 11 | Primary analysis | Ozturk 2009 |
| Australia West Caucasian | AUS-WCauc | Australia | Oceania | -31.950, 115.850 | 50 | 9 | Sensitivity analysis (diaspora / mixed ancestry) | Witt 1999 |
| England West Midlands Afro Caribbean | ENG-WM-AfC | Europe | Europe | 52.483, -1.900 | 54 | 11 | Sensitivity analysis (diaspora / mixed ancestry) | Cook 2003 |
| England West Midlands Indian Asian | ENG-WM-IndA | Europe | Europe | 52.483, -1.900 | 35 | 11 | Sensitivity analysis (diaspora / mixed ancestry) | Cook 2003 |
| Canada Iranian | CAN-Iranian | North America | North America | 49.250, -123.100 | 68 | 11 | Sensitivity analysis (diaspora / mixed ancestry) | Tavoularis |
| USA California African American | USA-CA-AfAm | North America | North America | 34.050, -118.250 | 58 | 11 | Sensitivity analysis (diaspora / mixed ancestry) | Du 2007 |
| USA California Asian American | USA-CA-AsAm | North America | North America | 34.050, -118.250 | 150 | 11 | Sensitivity analysis (diaspora / mixed ancestry) | Du 2007 |
| USA California Caucasians | USA-CA-Cauc | North America | North America | 34.050, -118.250 | 195 | 11 | Sensitivity analysis (diaspora / mixed ancestry) | Du 2007 |
| USA California Hispanic | USA-CA-Hisp | North America | North America | 34.050, -118.250 | 128 | 11 | Sensitivity analysis (diaspora / mixed ancestry) | Du 2007 |
| USA Caucasian | USA-Cauc | North America | North America | 38.883, -77.033 | 255 | 11 | Sensitivity analysis (diaspora / mixed ancestry) | Hollenbach 2010 |
| USA Springfield Puerto Rican | USA-PRican | North America | North America | 42.100, -72.533 | 74 | 11 | Sensitivity analysis (diaspora / mixed ancestry) | Zuniga 2006 |
| Brazil Belem Mixed | BRA-Belem | South and Central America | South America | -1.450, -48.500 | 347 | 11 | Sensitivity analysis (diaspora / mixed ancestry) | Pedroza 2011 |
| Brazil Belo Horizonte Caucasian | BRA-BH-Cauc | South and Central America | South America | -19.917, -43.917 | 90 | 11 | Sensitivity analysis (diaspora / mixed ancestry) | AFND only |
| Brazil Curitiba Japanese | BRA-Cur-Jpn | South and Central America | South America | -25.417, -49.267 | 51 | 10 | Sensitivity analysis (diaspora / mixed ancestry) | Augusto 2016 |
| Brazil Parana Japanese | BRA-Par-Jpn | South and Central America | South America | -22.433, -48.900 | 214 | 11 | Sensitivity analysis (diaspora / mixed ancestry) | AFND only |
| Brazil Parana Mixed | BRA-Par-Mx1 | South and Central America | South America | -24.000, -51.000 | 289 | 11 | Sensitivity analysis (diaspora / mixed ancestry) | Franceschi 2008 |
| Brazil Parana Mixed pop 2 | BRA-Par-Mx2 | South and Central America | South America | -24.000, -51.000 | 172 | 11 | Sensitivity analysis (diaspora / mixed ancestry) | Beltrame 2013 |
| Brazil Porto Alegre Caucasian | BRA-PA-Cau1 | South and Central America | South America | -30.017, -51.217 | 250 | 11 | Sensitivity analysis (diaspora / mixed ancestry) | Jobim 2008 |
| Brazil Porto Alegre Caucasian pop 2 | BRA-PA-Cau2 | South and Central America | South America | -30.017, -51.217 | 216 | 11 | Sensitivity analysis (diaspora / mixed ancestry) | Portela 2017 |
| Brazil Porto Alegre Caucasian pop 3 | BRA-PA-Cau3 | South and Central America | South America | -30.017, -51.217 | 278 | 11 | Sensitivity analysis (diaspora / mixed ancestry) | Jobim 2013 |
| Brazil Rio de Janeiro Caucasian pop 2 | BRA-RJ-Cauc | South and Central America | South America | -22.900, -43.183 | 98 | 11 | Sensitivity analysis (diaspora / mixed ancestry) | AFND only |
| Brazil Rio de Janeiro Mixed | BRA-RJ-Mix | South and Central America | South America | -22.883, -43.283 | 166 | 11 | Sensitivity analysis (diaspora / mixed ancestry) | Porto 2006 |
| Brazil Southeast Caucasian | BRA-SE-Cauc | South and Central America | South America | -23.000, -48.000 | 55 | 10 | Sensitivity analysis (diaspora / mixed ancestry) | Morgun 2004 |
| Cuba Caucasian | CUB-Cauc | South and Central America | Central America | 23.117, -82.367 | 70 | 11 | Sensitivity analysis (diaspora / mixed ancestry) | Middleton |
| Guadeloupe Mixed | GLP-Mixed | South and Central America | Central America | 15.983, -61.717 | 90 | 11 | Sensitivity analysis (diaspora / mixed ancestry) | Denis 2005 |
| Martinique Mixed | MTQ-Mixed | South and Central America | Central America | 14.667, -61.000 | 60 | 11 | Sensitivity analysis (diaspora / mixed ancestry) | Assaqa |
| Mexico Mestizo Guerrero | MEX-Mz-Guer | South and Central America | Central America | 17.550, -99.483 | 120 | 11 | Sensitivity analysis (diaspora / mixed ancestry) | Machado-Sulbaran 2017 |
| Mexico Mestizo Highlands | MEX-Mz-High | South and Central America | Central America | 19.433, -99.133 | 257 | 11 | Sensitivity analysis (diaspora / mixed ancestry) | AFND only |
| Mexico Mestizo Jalisco | MEX-Mz-Jal1 | South and Central America | Central America | 20.667, -103.350 | 100 | 11 | Sensitivity analysis (diaspora / mixed ancestry) | AFND only |
| Mexico Mestizo Jalisco pop 2 | MEX-Mz-Jal2 | South and Central America | Central America | 20.617, -103.333 | 124 | 11 | Sensitivity analysis (diaspora / mixed ancestry) | Machado-Sulbaran 2017 |
| Mexico Mestizo | MEX-Mz-1 | South and Central America | Central America | 19.417, -99.117 | 54 | 11 | Sensitivity analysis (diaspora / mixed ancestry) | Contreras 2007 |
| Mexico Mestizo pop 2 | MEX-Mz-2 | South and Central America | Central America | 24.483, -102.300 | 300 | 11 | Sensitivity analysis (diaspora / mixed ancestry) | AFND only |
| Mexico Mexico City Mestizo | MEX-Mz-City | South and Central America | Central America | 19.417, -99.117 | 86 | 11 | Sensitivity analysis (diaspora / mixed ancestry) | Gutierrez-Rodriguez 2006 |
| Mexico Torreon Mestizos | MEX-Mz-Torr | South and Central America | Central America | 25.533, -103.417 | 166 | 11 | Sensitivity analysis (diaspora / mixed ancestry) | AFND only |
| Mexico Veracruz Mestizo | MEX-Mz-Vera | South and Central America | Central America | 19.433, -96.367 | 51 | 11 | Sensitivity analysis (diaspora / mixed ancestry) | Middleton |
| Mexico West Mestizo | MEX-Mz-West | South and Central America | Central America | 29.083, -110.950 | 124 | 11 | Sensitivity analysis (diaspora / mixed ancestry) | AFND only |
| Trinidad Africans | TTO-African | South and Central America | Central America | 10.667, -61.517 | 62 | 11 | Sensitivity analysis (diaspora / mixed ancestry) | Norman 2002 |
| Trinidad South Asians | TTO-SAsian | South and Central America | Central America | 10.667, -61.517 | 108 | 11 | Sensitivity analysis (diaspora / mixed ancestry) | Norman 2002 |
| Uruguay Mixed | URY-Mixed | South and Central America | South America | -34.883, -56.167 | 41 | 11 | Sensitivity analysis (diaspora / mixed ancestry) | INDT Montevideo |
| Venezuela Mestizo | VEN-Mestizo | South and Central America | South America | 10.500, -66.917 | 205 | 11 | Sensitivity analysis (diaspora / mixed ancestry) | Fernandez-Mestre 2010 |
| Singapore Chinese | SGP-Chin1 | South-East Asia | Asia | 1.367, 103.800 | 47 | 11 | Sensitivity analysis (diaspora / mixed ancestry) | AFND only |
| Singapore Chinese pop 2 | SGP-Chin2 | South-East Asia | Asia | 1.367, 103.800 | 210 | 11 | Sensitivity analysis (diaspora / mixed ancestry) | Lee 2008 |
| Singapore Indians | SGP-Indian | South-East Asia | Asia | 1.367, 103.800 | 80 | 11 | Sensitivity analysis (diaspora / mixed ancestry) | Lee 2008 |
| Comoros Mixed | COM-Mixed | Sub-Saharan Africa | Africa | -11.683, 43.267 | 54 | 11 | Sensitivity analysis (diaspora / mixed ancestry) | Frassati 2006 |
| Reunion Mixed | REU-Mixed | Sub-Saharan Africa | Africa | -21.100, 55.517 | 101 | 11 | Sensitivity analysis (diaspora / mixed ancestry) | Denis 2005 |
| South Africa Caucasian | ZAF-Cauc | Sub-Saharan Africa | Africa | -26.133, 28.050 | 97 | 11 | Sensitivity analysis (diaspora / mixed ancestry) | Gentle 2017 |
| South Africa Indian | ZAF-Indian | Sub-Saharan Africa | Africa | -26.133, 28.050 | 50 | 11 | Sensitivity analysis (diaspora / mixed ancestry) | Gentle 2017 |
| South Africa Mixed Ancestry | ZAF-Mixed | Sub-Saharan Africa | Africa | -26.333, 28.050 | 50 | 11 | Sensitivity analysis (diaspora / mixed ancestry) | Gentle 2017 |

**Caption:** All 236 analysed populations, with the abbreviation used in Figure 2, geographic classification, coordinates, sample size, the number of variable KIR genes reported and the analysis in which each population was used. Abbreviations combine the ISO 3166-1 alpha-3 country code with a short locality or population descriptor where a country contributed more than one population. Populations labelled "Primary analysis" (n = 189) are geographically local. Populations labelled "Sensitivity analysis" (n = 47) were classified as recent diaspora, mixed-ancestry or Mestizo populations and were analysed only in the corresponding sensitivity analysis. A further 34 populations reporting fewer than 9 of the 11 variable genes were not analysed and are not listed. Coordinates are given as latitude, longitude in decimal degrees, with negative values south of the equator and west of the Greenwich meridian. The AFND population source publication is given as first author and year. **Abbreviations:** AFND: Allele Frequency Net Database; pop: population

**Supplementary Table 2.** Associations between gene-content measures and sampling effort.

| Measure | β on log sample size | SE | p | R² |
| --- | --- | --- | --- | --- |
| Expected heterozygosity | -0.0075 | 0.0077 | 0.3281 | 0.0051 |
| Standardized gene content | -0.1075 | 0.0697 | 0.1253 | 0.0126 |
| B-content | -0.0282 | 0.0119 | 0.0185 | 0.0293 |
| Activating index | -0.0166 | 0.0089 | 0.0647 | 0.0182 |
| Inhibitory index | -0.0009 | 0.0052 | 0.8642 | 0.0002 |

**Caption:** Each gene-content measure regressed on log-transformed sample size in the primary analysis set, fitted before any geographic comparison. Expected heterozygosity, standardized gene content and the inhibitory index showed no material association with sampling effort. B-content showed a weak negative association and the activating index a borderline association, and log sample size was therefore retained as a covariate in all subsequent regression models. **Abbreviations:** β: regression coefficient; SE: standard error; R²: coefficient of determination; He: expected heterozygosity.

**Supplementary Table 3.** Regional models of KIR diversity across analysis sets.

| Analysis set | Measure | Region F | p | Adjusted R² |
| --- | --- | --- | --- | --- |
| Primary analysis | Expected heterozygosity | 20.55 | < 0.0001 | 0.5347 |
| Primary analysis | Standardized gene content | 15.96 | < 0.0001 | 0.4721 |
| Sensitivity: n ≥ 50 | Expected heterozygosity | 18.88 | < 0.0001 | 0.5352 |
| Sensitivity: n ≥ 50 | Standardized gene content | 16.83 | < 0.0001 | 0.5137 |
| Sensitivity: expanded set | Expected heterozygosity | 16.73 | < 0.0001 | 0.4235 |
| Sensitivity: expanded set | Standardized gene content | 13.66 | < 0.0001 | 0.3748 |

**Caption:** Expected heterozygosity and standardized gene content modelled against geographic region with log-transformed sample size as a covariate, fitted separately in the primary analysis set and in each of the two sensitivity sets. Geographic region was strongly associated with both measures in every analysis set. **Abbreviations:** R²: coefficient of determination; He: expected heterozygosity.

**Supplementary Table 4.** Pairwise regional differentiation.

| Region 1 | Region 2 | n₁ | n₂ | R² | F | p | p (FDR) |
| --- | --- | --- | --- | --- | --- | --- | --- |
| Europe | North-East Asia | 39 | 32 | 0.5541 | 85.69 | 0.0001 | 0.0002 |
| Europe | South and Central America | 39 | 24 | 0.2882 | 24.69 | 0.0001 | 0.0002 |
| Europe | Western Asia | 39 | 19 | 0.1373 | 8.91 | 0.0001 | 0.0002 |
| Europe | South Asia | 39 | 17 | 0.2255 | 15.71 | 0.0001 | 0.0002 |
| Europe | Sub-Saharan Africa | 39 | 11 | 0.4876 | 45.66 | 0.0001 | 0.0002 |
| Europe | South-East Asia | 39 | 9 | 0.1474 | 7.93 | 0.0002 | 0.0003 |
| North-East Asia | South and Central America | 32 | 24 | 0.4278 | 40.24 | 0.0001 | 0.0002 |
| North-East Asia | Western Asia | 32 | 19 | 0.6051 | 75.03 | 0.0001 | 0.0002 |
| North-East Asia | Oceania | 32 | 6 | 0.3660 | 20.79 | 0.0001 | 0.0002 |
| North-East Asia | South Asia | 32 | 17 | 0.5393 | 54.99 | 0.0001 | 0.0002 |
| North-East Asia | Sub-Saharan Africa | 32 | 11 | 0.6301 | 69.78 | 0.0001 | 0.0002 |
| North-East Asia | South-East Asia | 32 | 9 | 0.1923 | 9.28 | 0.0001 | 0.0002 |
| South and Central America | South Asia | 24 | 17 | 0.1521 | 6.99 | 0.0001 | 0.0002 |
| South and Central America | Western Asia | 24 | 19 | 0.2516 | 13.77 | 0.0001 | 0.0002 |
| South and Central America | Sub-Saharan Africa | 24 | 11 | 0.4385 | 25.72 | 0.0001 | 0.0002 |
| Western Asia | Sub-Saharan Africa | 19 | 11 | 0.4686 | 24.64 | 0.0001 | 0.0002 |
| Sub-Saharan Africa | Oceania | 11 | 6 | 0.4061 | 10.26 | 0.0002 | 0.0003 |
| South Asia | South-East Asia | 17 | 9 | 0.2482 | 7.90 | 0.0020 | 0.0025 |
| South Asia | Sub-Saharan Africa | 17 | 11 | 0.3833 | 16.15 | 0.0001 | 0.0002 |
| Western Asia | South-East Asia | 19 | 9 | 0.3121 | 11.81 | 0.0001 | 0.0002 |
| Sub-Saharan Africa | South-East Asia | 11 | 9 | 0.5592 | 22.80 | 0.0001 | 0.0002 |
| South and Central America | South-East Asia | 24 | 9 | 0.1686 | 6.25 | 0.0020 | 0.0025 |
| Europe | Oceania | 39 | 6 | 0.1361 | 6.74 | 0.0079 | 0.0096 |
| South and Central America | Oceania | 24 | 6 | 0.1113 | 3.48 | 0.0140 | 0.0163 |
| Western Asia | South Asia | 19 | 17 | 0.0782 | 2.88 | 0.0292 | 0.0327 |
| Western Asia | Oceania | 19 | 6 | 0.0881 | 2.23 | 0.0709 | 0.0735 |
| South-East Asia | Oceania | 9 | 6 | 0.1962 | 3.16 | 0.0529 | 0.0570 |
| South Asia | Oceania | 17 | 6 | 0.0360 | 0.78 | 0.4710 | 0.4710 |

**Caption:** All 28 eligible pairwise comparisons between geographic regions represented by at least three complete profiles, from distance-based permutational analyses on Euclidean profile distances with 9,999 permutations. Twenty-five comparisons remained significant after Benjamini-Hochberg correction, applied as a step-up procedure with enforced monotonicity. Probability values of 0.0001 are at the resolution limit of 9,999 permutations. The same results are displayed in Supplementary Figure 2. **Abbreviations:** n₁, n₂: number of complete profiles in each region; R²: proportion of variation explained; F: pseudo-F statistic; FDR: false discovery rate.

**Supplementary Table 5.** Agreement among distance matrices and variance explained by the principal-coordinate axes.

|  | Value | Cumulative percent |
| --- | --- | --- |
| *A. Agreement among dissimilarity matrices (Mantel correlation, r)* | | |
| Euclidean vs Manhattan | 0.9761 | — |
| Euclidean vs Bray-Curtis | 0.9735 | — |
| Manhattan vs Bray-Curtis | 0.9898 | — |
| Euclidean vs z-scored Euclidean | 0.9646 | — |
| *B. Variance explained by the principal-coordinate axes (percent)* | | |
| PCo1 | 47.48 | 47.48 |
| PCo2 | 26.90 | 74.38 |
| PCo3 | 8.08 | 82.46 |
| PCo4 | 6.09 | 88.55 |
| PCo5 | 5.32 | 93.87 |
| PCo6 | 1.61 | 95.48 |

**Caption:** Both panels describe the same Euclidean ordination of the 161 complete 11-gene carrier-frequency profiles. Panel A gives Mantel correlations among the four dissimilarity matrices; Euclidean distance was the primary measure, and the high correlations indicate that the structural results do not depend on the choice of metric. Panel B gives the percentage of variation accounted for by each of the first six principal-coordinate axes of the Euclidean matrix, with the cumulative percentage. The first two axes account for 74.4% of the variation and are plotted in Figure 2. Cumulative percentages do not apply to the Mantel correlations and those cells are marked with an em dash. **Abbreviations:** r: Mantel correlation coefficient; PCo: principal coordinate.

**Supplementary Table 6.** Selection and regional agreement of the cluster solution.

| k | Mean silhouette width | ARI vs region | ARI vs broad area |
| --- | --- | --- | --- |
| 2 | 0.2925 | 0.1477 | 0.0619 |
| 3 | 0.3272 | 0.2363 | 0.1710 |
| 4 | 0.3254 | 0.3296 | 0.2721 |
| 5 | 0.3412 | 0.3427 | 0.2809 |
| 6 | 0.3558 | 0.3401 | 0.2762 |
| 7 | 0.3604 | 0.3315 | 0.2754 |
| 8 | 0.2230 | 0.3331 | 0.2850 |
| 9 | 0.2272 | 0.3322 | 0.2846 |
| 10 | 0.2177 | 0.3473 | 0.3019 |

**Caption:** Hierarchical clustering solutions containing 2 to 10 clusters, calculated on Euclidean distances among the 161 complete profiles using Ward’s minimum-variance method. Mean silhouette width is the internal criterion used to choose the number of clusters and reached its maximum at seven clusters, which was the solution carried forward. The two adjusted Rand indices measure agreement between each clustering solution and the external geographic classifications. They rise steeply from two to four clusters and then plateau, so agreement with geography is not what distinguishes the selected solution from its neighbours. Values well below 1 at every k indicate that the clusters capture broad geographic structure without corresponding to discrete regional groupings. Stability of the selected seven-cluster solution was assessed separately by resampling genes with replacement over 1,000 replicates and recording how frequently pairs of populations remained assigned to the same cluster: the median value for pairs grouped together in the full-data solution was 0.6519. Silhouette widths are plotted in Supplementary Figure 3. **Abbreviations:** k: number of clusters; ARI: adjusted Rand index.

**Supplementary Table 7.** Gene-specific differentiation among populations.

| Gene | G_ST_ | 95 % CI | |
| --- | --- | --- | --- |
|  |  | **LL** | **UL** |
| KIR2DS4 | 0.1065 | 0.0734 | 0.1356 |
| KIR2DL3 | 0.1045 | 0.0817 | 0.1258 |
| KIR2DL1 | 0.0972 | 0.0753 | 0.1209 |
| KIR3DL1 | 0.0738 | 0.0543 | 0.0959 |
| KIR2DL2 | 0.0652 | 0.0533 | 0.0776 |
| KIR2DS2 | 0.0596 | 0.0484 | 0.0704 |
| KIR2DS5 | 0.0555 | 0.0383 | 0.0760 |
| KIR3DS1 | 0.0505 | 0.0369 | 0.0639 |
| KIR2DS3 | 0.0486 | 0.0351 | 0.0627 |
| KIR2DS1 | 0.0454 | 0.0323 | 0.0610 |
| KIR2DL5 | 0.0445 | 0.0344 | 0.0539 |

**Caption:** Differentiation among the 161 complete-profile populations for each of the 11 variable KIR genes, quantified by Nei's coefficient of gene differentiation GST with each population treated as a subpopulation and no small-sample correction, calculated from presence-allele frequencies derived from carrier frequencies under Hardy-Weinberg equilibrium. Confidence intervals are percentile intervals from 1,000 bootstrap replicates obtained by resampling populations with replacement. The same estimates are plotted in Supplementary Figure 4. **Abbreviations:** G_ST_: coefficient of gene differentiation; CI: confidence interval; UL: upper limit; LL: lower limit.

**Supplementary Table 8.** KIR diversity and distance from East Africa.

| Outcome | Distance definition | β | SE | p | Adjusted R² | Clustered SE | Clustered p |
| --- | --- | --- | --- | --- | --- | --- | --- |
| Expected heterozygosity | waypoint | 0.0008 | 0.0005 | 0.1478 | 0.0057 | 0.0007 | 0.2463 |
| Expected heterozygosity | addis | -0.0028 | 0.0014 | 0.0552 | 0.0142 | 0.0026 | 0.2837 |
| Expected heterozygosity | nairobi | -0.0035 | 0.0016 | 0.0267 | 0.0207 | 0.0029 | 0.2320 |
| Expected heterozygosity | cairo | -0.0022 | 0.0013 | 0.0777 | 0.0112 | 0.0022 | 0.3028 |
| Standardized gene content | waypoint | 0.0018 | 0.0047 | 0.7006 | 0.0027 | 0.0067 | 0.7843 |
| Standardized gene content | addis | -0.0378 | 0.0129 | 0.0038 | 0.0460 | 0.0226 | 0.0941 |
| Standardized gene content | nairobi | -0.0470 | 0.0139 | 0.0008 | 0.0602 | 0.0253 | 0.0628 |
| Standardized gene content | cairo | -0.0298 | 0.0114 | 0.0096 | 0.0374 | 0.0193 | 0.1218 |

**Caption:** Expected heterozygosity and standardized gene content regressed on distance from East Africa under four distance definitions, each adjusted for log-transformed sample size. Coefficients are expressed per 1,000 km. The waypoint-constrained route was the prespecified primary definition. The prespecified waypoint-constrained route gave no decline, whereas all three great-circle definitions gave negative coefficients. Standard errors and probability values are from ordinary least squares; the final two columns give country-clustered standard errors and the corresponding probability values, which treat populations sampled within one country as non-independent. The primary relationship is plotted in Supplementary Figure 5. **Abbreviations:** β: regression coefficient; SE: standard error; R²: coefficient of determination; He: expected heterozygosity

**Supplementary Table 9.** Distance from East Africa across all populations and restricted to the Old World

| Outcome | Distance definition | All populations β | p | Old World β | p | Clustered p, all populations | Clustered p, Old World | Interaction p (clustered) |
| --- | --- | --- | --- | --- | --- | --- | --- | --- |
| Expected heterozygosity | Waypoint-constrained | 0.0008 | 0.1478 | -0.0066 | 0.0001 | 0.2463 | 0.0823 | 0.0085 (0.1428) |
| Expected heterozygosity | Addis Ababa great-circle | -0.0028 | 0.0552 | -0.0158 | < 0.0001 | 0.2837 | 0.0001 | 0.3079 (0.4055) |
| Expected heterozygosity | Nairobi great-circle | -0.0035 | 0.0267 | -0.0143 | < 0.0001 | 0.2320 | 0.0015 | 0.3710 (0.4932) |
| Expected heterozygosity | Cairo great-circle | -0.0022 | 0.0777 | -0.0119 | < 0.0001 | 0.3028 | 0.0013 | 0.1635 (0.1667) |
| Standardized gene content | Waypoint-constrained | 0.0018 | 0.7006 | -0.0789 | < 0.0001 | 0.7843 | 0.0092 | 0.0005 (0.0426) |
| Standardized gene content | Addis Ababa great-circle | -0.0378 | 0.0038 | -0.1615 | < 0.0001 | 0.0941 | 0.0000 | 0.1073 (0.1170) |
| Standardized gene content | Nairobi great-circle | -0.0470 | 0.0008 | -0.1563 | < 0.0001 | 0.0628 | 0.0000 | 0.0432 (0.0828) |
| Standardized gene content | Cairo great-circle | -0.0298 | 0.0096 | -0.1132 | < 0.0001 | 0.1218 | 0.0004 | 0.3115 (0.3686) |

**Caption:** Expected heterozygosity and standardized gene content regressed on distance from East Africa under the four distance definitions, adjusted for log-transformed sample size, fitted across all 189 populations and across the 155 populations outside the Americas, Oceania and Australia. Coefficients are per 1,000 km. Across all populations no definition shows a decline under the prespecified waypoint route. Restricted to the Old World, every definition shows a steep and significant decline in both measures. The interaction column gives the ordinary-least-squares probability value with the country-clustered value in parentheses. Clustered probability values for the all-population and Old World models are also given. Clustering widens every interval; the decline in standardized gene content across the Old World and its interaction with the New World remain significant under clustering, whereas the corresponding expected-heterozygosity results do not. **Abbreviations:** β: regression coefficient.

**Supplementary Table 10.** Global latitude models for absolute and signed latitude.

| Outcome | Absolute latitude | | | | | Signed latitude | | | | |
| --- | --- | --- | --- | --- | --- | --- | --- | --- | --- | --- |
|  | **β** | **SE** | **p** | **Clustered p** | **Adjusted R²** | **β** | **SE** | **p** | **Clustered p** | **Adjusted R²** |
| Expected heterozygosity | -0.0004 | 0.0003 | 0.2450 | 0.3766 | 0.0017 | -0.0004 | 0.0002 | 0.0660 | 0.1099 | 0.0126 |
| Standardized gene content | -0.0030 | 0.0030 | 0.3289 | 0.4399 | 0.0071 | -0.0033 | 0.0018 | 0.0615 | 0.0788 | 0.0206 |
| B-content | -0.0010 | 0.0005 | 0.0419 | 0.1143 | 0.0405 | -0.0009 | 0.0003 | 0.0021 | 0.0041 | 0.0675 |
| B-proportion | -0.0012 | 0.0006 | 0.0298 | 0.0798 | 0.0456 | -0.0011 | 0.0003 | 0.0006 | 0.0009 | 0.0805 |
| Activating index | -0.0004 | 0.0004 | 0.2451 | 0.3836 | 0.0149 | -0.0005 | 0.0002 | 0.0273 | 0.0753 | 0.0334 |
| Inhibitory index | 0.0001 | 0.0002 | 0.5271 | 0.5191 | -0.0084 | 0.0001 | 0.0001 | 0.6908 | 0.7477 | -0.0097 |

**Caption:** Each of the six gene-content measures regressed on latitude in the primary analysis set (n = 189), adjusted for log-transformed sample size. Absolute latitude represents distance from the equator and was the prespecified primary measure; signed latitude tests directional north-south variation and is reported here as the corresponding secondary specification. Coefficients are expressed per degree of latitude. B-content and B-proportion declined with both measures, and the signed-latitude models were the more strongly supported of the two. The absolute-latitude relationship for B-content is plotted in Figure 3. The final two columns give probability values from country-clustered standard errors. **Abbreviations:** n: number of populations; β: regression coefficient; SE: standard error; R²: coefficient of determination; He: expected heterozygosity

**Supplementary Table 11.** Within-region absolute-latitude models.

| Region | Outcome | n | β per degree | SE | p | p (FDR) |
| --- | --- | --- | --- | --- | --- | --- |
| Europe | B-content | 52 | -0.0023 | 0.0009 | 0.0112 | 0.0784 |
| Europe | Inhibitory index | 52 | -0.0007 | 0.0007 | 0.3433 | 0.4005 |
| North-East Asia | B-content | 35 | -0.0001 | 0.0016 | 0.9611 | 0.9611 |
| North-East Asia | Inhibitory index | 35 | -0.0009 | 0.0008 | 0.2694 | 0.4005 |
| South and Central America | B-content | 25 | -0.0033 | 0.0017 | 0.0700 | 0.1633 |
| South and Central America | Inhibitory index | 25 | -0.0002 | 0.0006 | 0.7213 | 0.7213 |
| Western Asia | B-content | 22 | 0.0001 | 0.0013 | 0.9444 | 0.9611 |
| Western Asia | Inhibitory index | 22 | -0.0013 | 0.0010 | 0.1983 | 0.4005 |
| South Asia | B-content | 18 | -0.0077 | 0.0050 | 0.1412 | 0.2471 |
| South Asia | Inhibitory index | 18 | -0.0033 | 0.0016 | 0.0521 | 0.3647 |
| South-East Asia | B-content | 14 | -0.0028 | 0.0046 | 0.5545 | 0.7763 |
| South-East Asia | Inhibitory index | 14 | 0.0016 | 0.0016 | 0.3426 | 0.4005 |
| Sub-Saharan Africa | B-content | 11 | 0.0023 | 0.0011 | 0.0634 | 0.1633 |
| Sub-Saharan Africa | Inhibitory index | 11 | 0.0007 | 0.0006 | 0.2441 | 0.4005 |

**Caption:** B-content and the inhibitory index regressed on absolute latitude separately within each geographic region represented by at least 10 populations, adjusted for log-transformed sample size. Probability values were adjusted separately by outcome using the Benjamini-Hochberg step-up procedure with enforced monotonicity. No within-region association remained significant after correction. The B-content coefficients are plotted in Supplementary Figure 6. **Abbreviations:** n: number of populations; β: regression coefficient; SE: standard error; FDR: false discovery rate.

**Supplementary Table 12.** Joint latitude and distance models.

| Outcome | Model | Term | β | SE | p | Clustered p |
| --- | --- | --- | --- | --- | --- | --- |
| B-content | latitude only | absolute latitude | -0.0010 | 0.0005 | 0.0419 | 0.1143 |
| B-content | distance only | waypoint distance | 0.0010 | 0.0008 | 0.1412 | 0.2160 |
| B-content | joint | absolute latitude | -0.0009 | 0.0005 | 0.1078 | 0.2597 |
| B-content | joint | waypoint distance | 0.0007 | 0.0009 | 0.4288 | 0.5553 |
| Expected heterozygosity | latitude only | absolute latitude | -0.0004 | 0.0003 | 0.2450 | 0.3766 |
| Expected heterozygosity | distance only | waypoint distance | 0.0008 | 0.0005 | 0.1478 | 0.2463 |
| Expected heterozygosity | joint | absolute latitude | -0.0002 | 0.0004 | 0.4944 | 0.6399 |
| Expected heterozygosity | joint | waypoint distance | 0.0006 | 0.0006 | 0.2721 | 0.4373 |

**Caption:** B-content and expected heterozygosity modelled against absolute latitude alone, waypoint-constrained distance from East Africa alone, and both predictors together, each adjusted for log-transformed sample size. The joint models test whether either predictor retains an independent association. The final column gives probability values from country-clustered standard errors. **Abbreviations**: β: regression coefficient; SE: standard error; He: expected heterozygosity

**Supplementary Table 13.** Simulation-based power analyses.

| Outcome | Predictor | Effect size | Power |
| --- | --- | --- | --- |
| B-content | Absolute latitude | 0.0002 | 0.0582 |
| B-content | Absolute latitude | 0.0005 | 0.1481 |
| B-content | Absolute latitude | 0.0010 | 0.5340 |
| B-content | Absolute latitude | 0.0015 | 0.8363 |
| B-content | Absolute latitude | 0.0020 | 0.9741 |
| B-content | Absolute latitude | 0.0030 | 1.0000 |
| Expected heterozygosity | Waypoint distance | 0.0005 | 0.1761 |
| Expected heterozygosity | Waypoint distance | 0.0010 | 0.4466 |
| Expected heterozygosity | Waypoint distance | 0.0020 | 0.9684 |
| Expected heterozygosity | Waypoint distance | 0.0030 | 1.0000 |
| Expected heterozygosity | Waypoint distance | 0.0050 | 1.0000 |

**Caption:** Probability of detecting prespecified effects of absolute latitude on B-content and of waypoint-constrained distance on expected heterozygosity, estimated by simulation conditional on the observed sample size, predictor distributions, association with log-transformed sample size and residual variance. Effect sizes are expressed per degree of latitude and per 1,000 km respectively, with 1,000 simulations per effect size. The curves are plotted in Supplementary Figure 9, panel B. **Abbreviations:** He: expected heterozygosity

**Supplementary Table 14.** Latitude models under precision weighting.

| Outcome | Unweighted β | p | Weight = n β | p | Inverse-variance β | p |
| --- | --- | --- | --- | --- | --- | --- |
| Expected heterozygosity | -0.0004 | 0.2450 | 0.0001 | 0.8216 | 0.0001 | 0.6779 |
| Standardized gene content | -0.0030 | 0.3289 | -0.0006 | 0.8320 | -0.0003 | 0.9177 |
| B-content | -0.0010 | 0.0419 | -0.0004 | 0.3638 | -0.0004 | 0.4553 |
| B-proportion | -0.0012 | 0.0298 | -0.0005 | 0.3847 | -0.0004 | 0.4871 |
| Activating index | -0.0004 | 0.2451 | -0.0000 | 0.8971 | -0.0000 | 0.9855 |
| Inhibitory index | 0.0001 | 0.5271 | 0.0001 | 0.7155 | 0.0001 | 0.6378 |

**Caption:** Each gene-content measure regressed on absolute latitude in the primary analysis set (n = 189), adjusted for log-transformed sample size, under three weighting schemes. Sample sizes range from 19 to 690 individuals, and the unweighted models give every population equal influence. Weights proportional to sample size and weights proportional to the inverse of the binomial sampling variance of the outcome both remove the B-content and B-proportion associations with absolute latitude, indicating that those associations rest disproportionately on populations estimated with the least precision. Coefficients are expressed per degree of latitude. **Abbreviations:** β: regression coefficient.

**Supplementary Table 15.** Latitude and distance models refitted as spatial models.

| Model or predictor | β | SE | p | Residual Moran's I |
| --- | --- | --- | --- | --- |
| Ordinary least squares | -0.0010 | 0.0005 | 0.0419 | 0.4387 |
| GLS, exponential correlation  (range 3,090 km, nugget 0.273) | -0.0011 | 0.0011 | 0.3343 | — |
| Moran eigenvector filtering (6 eigenvectors) | -0.0001 | 0.0005 | 0.7875 | 0.0001 |
| Collinearity with region: absolute latitude | η² = 0.7627 | — | — | — |
| Collinearity with region: signed latitude | η² = 0.8343 | — | — | — |
| Collinearity with region: waypoint distance | η² = 0.9788 | — | — | — |
| Collinearity with region: Addis Ababa distance | η² = 0.8879 | — | — | — |
| Collinearity with region: Nairobi distance | η² = 0.8537 | — | — | — |
| Collinearity with region: Cairo distance | η² = 0.9292 | — | — | — |

**Caption:** B-content regressed on absolute latitude with log-transformed sample size, fitted by ordinary least squares and by two spatial specifications. Generalised least squares used an exponential correlation structure on great-circle distance, with the range and nugget estimated by maximum likelihood. Moran eigenvector filtering added spatial eigenvectors selected to minimise residual autocorrelation. The table also gives the collinearity of each continuous geographic predictor with geographic region, as the proportion of its variance explained by the 12 regions. Under either spatial specification the latitude association is no longer supported. The residual Moran’s I for the ordinary-least-squares model is the value reported in Supplementary Table 17. **Abbreviations:** β: regression coefficient; GLS: generalised least squares; η²: proportion of variance explained by region

**Supplementary Table 16.** Shape of the latitude relationship: hemisphere interaction and quadratic specification.

| Outcome | Latitude × hemisphere interaction | | | Quadratic absolute latitude | | | | |
| --- | --- | --- | --- | --- | --- | --- | --- | --- |
|  | **β** | **SE** | **p** | **β**  **(× 10⁻⁵)** | **p** | **Linear AIC** | **Quadratic AIC** | **Nested p** |
| Expected heterozygosity | 0.0003 | 0.0011 | 0.7570 | 3.20 | 0.0861 | -498.7 | -499.7 | 0.0860 |
| Standardized gene content | -0.0048 | 0.0098 | 0.6211 | 15.84 | 0.3522 | 335.9 | 337.0 | 0.3522 |
| B-content | -0.0008 | 0.0016 | 0.6103 | 3.23 | 0.2613 | -336.3 | -335.6 | 0.2611 |
| B-proportion | -0.0008 | 0.0018 | 0.6264 | 4.32 | 0.1731 | -298.6 | -298.5 | 0.1730 |
| Activating index | -0.0004 | 0.0012 | 0.7021 | 1.10 | 0.6120 | -443.0 | -441.2 | 0.6123 |
| Inhibitory index | -0.0003 | 0.0007 | 0.6806 | 1.58 | 0.2162 | -643.4 | -642.9 | 0.2165 |

**Caption:** Two specifications testing the shape of the absolute-latitude relationship, each fitted for all six gene-content measures with log-transformed sample size as a covariate and compared with the linear absolute-latitude models in Supplementary Table 10. The interaction term tests whether the absolute-latitude slope differs between the northern and southern hemispheres, and is coded so that a positive coefficient indicates a steeper slope in the northern hemisphere; no interaction approached statistical significance. The quadratic term tests departure from linearity and was compared with the linear specification using information criteria and a nested-model test; it did not improve fit for any outcome by the nested-model test, and the only outcome whose Akaike information criterion fell was expected heterozygosity, by 1.0 unit, which is not material support for a quadratic term. Interaction coefficients are expressed per degree of latitude and quadratic coefficients per squared degree, the latter multiplied by 10⁻⁵ for legibility. The coefficients are plotted in Supplementary Figure 7, panels A and B respectively. **Abbreviations:** β: regression coefficient; SE: standard error; AIC: Akaike information criterion.

**Supplementary Table 17.** Residual spatial autocorrelation.

| Model | n | Moran's I | Permutation p |
| --- | --- | --- | --- |
| B-content ~ absolute latitude | 189 | 0.4387 | 0.0010 |
| He ~ waypoint distance | 189 | 0.4507 | 0.0010 |
| B-content joint model | 189 | 0.4415 | 0.0010 |
| He joint model | 189 | 0.4564 | 0.0010 |

**Caption:** Moran’s I calculated for residuals from the primary latitude, distance and joint models, using a row-standardized eight-nearest-neighbour spatial network constructed from population coordinates. Significant residual spatial autocorrelation indicates that conventional non-spatial standard errors and probability values are anti-conservative. **Abbreviations**: I: Moran’s I; n: number of populations; He: expected heterozygosity.

**Supplementary Table 18.** Multivariate-outlier screen.

| Population | Region | Mahalanobis distance | Raw p | p (FDR) | Outlier |
| --- | --- | --- | --- | --- | --- |
| India North pop 2 | South Asia | 66.50 | < 0.0001 | < 0.0001 | Yes |
| China Hubei Province Tujia | North-East Asia | 65.63 | < 0.0001 | < 0.0001 | Yes |
| Papua New Guinea Irian Jaya | Oceania | 60.72 | < 0.0001 | < 0.0001 | Yes |
| India Northeast Assam Adivasi | South Asia | 45.09 | < 0.0001 | < 0.0001 | Yes |
| Costa Rica Guanacaste | South and Central America | 45.00 | < 0.0001 | < 0.0001 | Yes |
| Australia Queensland Brisbane | Australia | 44.02 | < 0.0001 | < 0.0001 | Yes |
| Georgia | Western Asia | 37.53 | < 0.0001 | 0.0021 | Yes |
| Venezuela Yucpa | South and Central America | 37.52 | < 0.0001 | 0.0020 | Yes |
| Uzbekistan | Central Asia | 34.76 | < 0.0001 | 0.0051 | Yes |
| China Inner Mongolia Province pop 2 | North-East Asia | 31.97 | < 0.0001 | 0.0122 | Yes |
| Chile Talca | South and Central America | 30.88 | 0.0010 | 0.0171 | Yes |
| Brazil Amazon | South and Central America | 30.38 | 0.0012 | 0.0181 | Yes |
| China Xinjiang Region Urumqi Uygur | North-East Asia | 30.22 | 0.0020 | 0.0182 | Yes |
| South Africa San | Sub-Saharan Africa | 27.50 | 0.0041 | 0.0441 | Yes |
| USA Eastern | North America | 26.71 | 0.0051 | 0.0526 | No |
| India North Hindu | South Asia | 26.68 | 0.0056 | 0.0522 | No |
| South Africa Xhosa | Sub-Saharan Africa | 25.31 | 0.0081 | 0.0744 | No |
| China Yunnan Province Menghai Bulang | North-East Asia | 25.28 | 0.0080 | 0.0740 | No |
| Argentina Salta Wichi | South and Central America | 24.54 | 0.0111 | 0.0903 | No |
| Mexico Tarahumara | South and Central America | 23.45 | 0.0152 | 0.1235 | No |
| East Timor | South-East Asia | 23.11 | 0.0171 | 0.1317 | No |
| India North | South Asia | 22.69 | 0.0206 | 0.1431 | No |
| Solomon Islands | Oceania | 22.38 | 0.0225 | 0.1512 | No |
| Ivory Coast Abidjan | Sub-Saharan Africa | 21.20 | 0.0311 | 0.2106 | No |
| Portugal Azores Terceira Island | Europe | 20.82 | 0.0352 | 0.2274 | No |
| Tonga | Oceania | 19.55 | 0.0525 | 0.3222 | No |
| Indonesia Java | South-East Asia | 18.67 | 0.0674 | 0.4011 | No |
| Mexico Purepecha | South and Central America | 17.97 | 0.0823 | 0.4730 | No |
| Macedonia Romani | Europe | 17.09 | 0.1051 | 0.5848 | No |
| Brazil Guarani Kaiowá | South and Central America | 16.67 | 0.1182 | 0.6336 | No |
| Ghana | Sub-Saharan Africa | 16.50 | 0.1241 | 0.6424 | No |
| Borneo Kalimantan | South-East Asia | 16.37 | 0.1286 | 0.6426 | No |
| India Tamil Nadu Kanikar | South Asia | 16.26 | 0.1325 | 0.6423 | No |
| Brazil Kaingang | South and Central America | 15.66 | 0.1541 | 0.7301 | No |
| Pakistan Karachi | South Asia | 15.24 | 0.1721 | 0.7581 | No |
| Zimbabwe Harare | Sub-Saharan Africa | 15.20 | 0.1733 | 0.7580 | No |
| Tokelau | Oceania | 15.19 | 0.1745 | 0.7580 | No |
| Taiwan Han pop 4 | North-East Asia | 14.74 | 0.1954 | 0.8253 | No |
| Argentina Chaco Region Wichi | South and Central America | 14.04 | 0.2316 | 0.9341 | No |
| Latvia | Europe | 13.98 | 0.2341 | 0.9344 | No |

**Caption:** The 40 most extreme populations on a Mahalanobis-distance screen of the 161 complete 11-gene profiles. Raw probabilities are from a chi-square distribution of 11 degrees of freedom and were adjusted using the Benjamini-Hochberg procedure. Populations classified as outliers were retained in all primary analyses and excluded only in the corresponding sensitivity analysis. **Abbreviations**: FDR: false discovery rate

**Supplementary Table 19.** Leave-one-region-out analyses.

| Dropped region | n | Regional R² | p |
| --- | --- | --- | --- |
| South and Central America | 137 | 0.5762 | 0.0021 |
| Australia | 160 | 0.5281 | 0.0023 |
| South-East Asia | 152 | 0.5433 | 0.0022 |
| North-East Asia | 129 | 0.4124 | 0.0021 |
| Oceania | 155 | 0.5526 | 0.0020 |
| Europe | 122 | 0.5475 | 0.0020 |
| Sub-Saharan Africa | 150 | 0.4962 | 0.0023 |
| Western Asia | 142 | 0.5363 | 0.0020 |
| South Asia | 144 | 0.5581 | 0.0025 |
| North Africa | 160 | 0.5303 | 0.0021 |
| North America | 160 | 0.5317 | 0.0023 |
| Central Asia | 160 | 0.5333 | 0.0027 |

**Caption**: Proportion of multivariate variation attributable to geographic region after omitting each region in turn, from distance-based permutational analyses. The full-data value was 0.5349. Omitting North-East Asia reduced the regional R² to 0.412, whereas omitting any other region changed it by less than 0.05 (the largest, for South and Central America, was 0.0411), indicating that a substantial part of the regional signal derives from the contrast between East Asia and the remaining regions. The values are plotted in Supplementary Figure 9, panel A. **Abbreviations:** n: number of complete profiles retained; R²: proportion of variation explained.

**Supplementary Table 20.** Variance decomposition against a permutation null, and robustness of the regional component.

| Analysis | df | R² | Adjusted R² | Permutation null R² | p | p, publication year | p, typing method |
| --- | --- | --- | --- | --- | --- | --- | --- |
| Region | 11 | 0.5349 | 0.5005 | 0.0688 | 0.0005 | — | — |
| Country within region | 51 | 0.1878 | — | 0.3181 | 1.0000 | — | — |
| Residual | 98 | 0.2773 | — | — | — | — | — |
| Region and country combined | 62 | 0.7227 | 0.5472 | — | — | — | — |
| Finer East Asian partition (14 groups) | 13 | 0.5503 | 0.5106 | — | 0.0010 | — | — |
| Shared sampling locations collapsed (145 profiles) | 11 | 0.5420 | — | — | — | — | — |
| Subset with typing metadata (151 profiles) | 11 | 0.5462 | — | — | — | — | — |
| Region adjusted for year and typing method, expected heterozygosity | 11 | — | 0.5482 | — | < 0.0001 | 0.7991 | 0.1544 |
| Region adjusted for year and typing method, gene content | 11 | — | 0.4861 | — | < 0.0001 | 0.8675 | 0.1916 |

**Caption:** The upper rows refer each variance component to the distribution obtained by permuting the same grouping within the same unbalanced design, with degrees of freedom and adjusted R². The regional component lies far above its null. The country-within-region component lies below its null and therefore reflects the 51 degrees of freedom it consumes across 63 countries, 38 of which contribute a single profile, rather than explained variation. The lower rows refit the regional model under each dependency identified in review. The final two columns give the probability values for publication year and for typing method in the two adjusted models. **Abbreviations:** R²: proportion of variation explained

**Supplementary Table 21.** Carrier frequency by typing method and publication year, by gene.

| Gene | Missing pre-2010 (%) | Missing 2010 onwards (%) | β for SSP-only typing | p | p for publication year |
| --- | --- | --- | --- | --- | --- |
| KIR2DL1 | 0.9 | 0.0 | -0.0073 | 0.4796 | 0.0323 |
| KIR2DL2 | 0.0 | 0.0 | -0.0598 | 0.0428 | 0.7206 |
| KIR2DL3 | 0.0 | 0.0 | +0.0392 | 0.0096 | 0.7069 |
| KIR2DL5 | 16.2 | 6.0 | -0.0242 | 0.2991 | 0.6012 |
| KIR3DL1 | 0.9 | 0.0 | -0.0133 | 0.3206 | 0.0769 |
| KIR2DS1 | 0.0 | 0.0 | +0.0346 | 0.1221 | 0.3530 |
| KIR2DS2 | 0.0 | 0.0 | -0.0675 | 0.0171 | 0.4058 |
| KIR2DS3 | 0.9 | 0.0 | -0.0232 | 0.3370 | 0.3167 |
| KIR2DS4 | 3.6 | 3.0 | -0.0359 | 0.0705 | 0.2253 |
| KIR2DS5 | 3.6 | 0.0 | -0.0024 | 0.9177 | 0.5900 |
| KIR3DS1 | 0.0 | 1.5 | +0.0343 | 0.1547 | 0.3368 |

**Caption:** Each gene's carrier frequency regressed on an indicator for sequence-specific primer typing without sequence-specific oligonucleotide probe confirmation, and on publication year, across the 178 primary-set populations whose database record gives both. Panel completeness is reported for each era. KIR2DL5 is the gene most often absent from older panels. Publication year was associated with carrier frequency for KIR2DL1 only (p = 0.0323), which is the single nominally significant year effect across the 11 genes and is not corrected for multiple testing. Carrier frequencies differ by typing method for KIR2DL2, KIR2DL3 and KIR2DS2, the centromeric cluster containing one allelic pair, and not for KIR2DS4, KIR2DL5, KIR2DS3 or KIR2DS5, for which method-dependent false negatives have been described. **Abbreviations:** β: regression coefficient; SSP: sequence-specific primer.

**Supplementary Table 22.** Regional differentiation after excluding sparsely represented regions and at stricter sample-size thresholds.

| Analysis | Profiles | Regions | R² | F | p |
| --- | --- | --- | --- | --- | --- |
| Primary analysis, all 12 regions | 161 | 12 | 0.5349 | 15.58 | 0.0001 |
| Four sparse regions excluded | 157 | 8 | 0.5181 | 22.89 | 0.0010 |
| Four sparse regions excluded, n ≥ 50 | 142 | 8 | 0.5363 | 22.14 | 0.0010 |
| Four sparse regions excluded, n ≥ 100 | 97 | 7 | 0.5442 | 17.91 | 0.0010 |
| Four sparse regions excluded, n ≥ 150 | 46 | 7 | 0.7294 | 17.52 | 0.0010 |

**Caption:** Distance-based permutational analysis of geographic region among complete 11-gene profiles. The first row is the primary analysis. The remaining rows exclude the four regions represented by fewer than three complete profiles (Central Asia, North America, Australia and North Africa), which contribute degrees of freedom without informing within-region variation, and then restrict the remaining profiles to progressively larger samples. Because distance-based permutational analysis does not accommodate case weights, these thresholds serve as the analogue of precision weighting for the multivariate models. **Abbreviations:** R²: proportion of variation explained; F: pseudo-F statistic.
